# A nanobody-functionalized organic electrochemical transistor for the rapid detection of SARS-CoV-2 and MERS antigens at the physical limit

**DOI:** 10.1101/2020.11.12.20228874

**Authors:** Keying Guo, Shofarul Wustoni, Anil Koklu, Escarlet Díaz-Galicia, Maximilian Moser, Adel Hama, Ahmed A. Alqahtani, Adeel Nazir Ahmad, Fatimah Saeed Alhamlan, Iain McCulloch, Stefan T. Arold, Raik Grünberg, Sahika Inal

## Abstract

The COVID-19 pandemic highlights the need for rapid protein detection and quantification at the single-molecule level in a format that is simple and robust enough for widespread point-of-care applications. We here introduce a modular nanobody-organic electrochemical transistor architecture that enables the fast and specific detection and quantification of single-molecule to nanomolar protein antigen concentrations in complex bodily fluids. The sensor combines a new solution-processable organic semiconductor material in the transistor channel with the high-density and orientation-controlled bioconjugation of nanobody fusion proteins on disposable gate electrodes. It provides results after a 10 minutes exposure to 5 µL of unprocessed samples, maintains high specificity and single-molecule sensitivity in human saliva or serum, and is rapidly reprogrammed towards any protein target for which nanobodies exist. We demonstrate the use of this highly modular platform for the detection of green fluorescent protein, SARS-CoV-1/2, and MERS-CoV spike proteins and validate the sensor for COVID-19 screening in unprocessed clinical nasopharyngeal swab and saliva samples.

---

SARS-CoV-2 has caused more than one million fatalities as of October 2020^1^, and may remain a worldwide health and economic burden for several years^2^. Owing to airborne transmission^3^ and a large share of symptom-free but infective carriers^4,5^, this virus defies traditional suppression measures that are based on the isolation of symptomatic individuals. Molecular testing has therefore become an essential tool in global attempts to curb the COVID-19 pandemic^6^. Testing currently relies on the Reverse Transcription Polymerase Chain Reaction (RT-PCR), a well-established diagnostic method with close to single-molecule sensitivity^7^. RT-PCR, however, requires complex sample processing in specialized, well-instrumented laboratories. Due to the limited capacity of these laboratories, and the required logistics, results are typically communicated one to several days after sample collection. This delay in isolating infected carriers is severely hampering the suppression of infection chains^8^.

In a point-of-care (POC) setting, lateral flow immunoassays could offer an alternative as they detect viral protein (rather than RNA) directly in unprocessed patient samples and give qualitative results within 15 to 30 min. Yet, fast immunoassays detect viral loads above 50 million copies/mL, which is useful for the isolation of highly infectious individuals, but not sensitive enough for diagnostic purposes^9^. Enzymatic nucleic acid detection based POC assays, on the other hand, generally have to trade off sensitivity and robustness against sample processing and reaction times^10^. Thus, the COVID-19 crisis highlights a critical limitation of our current POC diagnostics toolbox. What has remained elusive so far is rapid protein detection at the single-molecule level in a format that is simple and robust enough for widespread POC or bedside application. We here present a bioelectronic sensor platform that fulfils these requirements.

Recent years have seen major progress in the development of transistors which transduce and, importantly, amplify biological reactions or binding events into an electrical readout^11–14^. Organic electrochemical transistors (OECTs) have emerged as an alternative bioelectronic transducer that outperforms all other electrolyte gated transistors and performs favorably compared to most solid-state technologies with their record-high transconductance values^15^. OECTs operate in aqueous media and integrate ion-permeable conjugated polymers in their channel which is volumetrically (de)doped by electrolyte ions injected by a gate electrode. The volumetric coupling between ionic and electronic charges in the channel makes OECTs powerful voltage amplifiers, as such, even a few binding events at the gate electrode can cause large modulations in the channel current^16–18^. This efficient, on-site, amplification of input signal allows for miniaturization and low-noise recordings, which should make OECTs particularly suitable for POC applications^17^. Despite these advantages, there are only few reports of label-free OECT immunosensors^19,20^. More generally, current electronic protein sensors appear to translate poorly into real-world applications. Common drawbacks include complex sensor designs, high noise, the need for tightly controlled environments, reliance on regular off-the-shelf antibodies and their chemical immobilization in random orientations^21^.

We overcame these limitations in protein detection with a bioelectronic platform that advances OECT technology on several levels, ranging from chemical materials to biological recognition. Key features in this process were (i) the use of a novel organic semiconductor that allowed for the first practical implementation of an accumulation-mode OECT and (ii) the controlled biofunctionalization of the sensor surface with (iii) custom-engineered nanobodies^22^. The accumulation-mode OECT combined high output signal strength and stability with very low input power requirement. The bioassembly strategy coupled recombinant protein in a precisely defined orientation at very high physical density.

We demonstrate that the resulting platform is fast (<15 min from sample to result) and works with unprocessed saliva or serum samples. The sensor is largely reusable, easy to manufacture, and highly modular. It reliably and specifically detects single protein molecules on millimeter-sized disposable electrodes in uncontrolled, ambient environments. The sensor is also quantitative and distinguishes concentrations over 8 to 10 orders of magnitude (attomolar to nanomolar). Experience with three different target proteins shows that our platform is broadly applicable and only limited by the availability of antigen-specific nanobodies. We validated the performance with clinical unprocessed nasopharyngeal swab and saliva samples from COVID-19 patients, and demonstrate a sensitivity comparable to RT-PCR methods.

## Biofunctionalization

Nanobodies^22^ are compact recognition modules made from the antigen-binding domain (VHH) of an unusual class of heavy chain-only antibodies found in Camelids^23^. Although several therapeutic^24^ and diagnostic applications^25^ have been developed, nanobodies have not yet been combined with OECT technology. One report described the functionalization of a conventional field effect transistor (FET) with nanobodies. This platform relied on random chemical immobilization of nanobodies on carbon nanotubes and reached ∼1 pM detection limit for green fluorescent protein (GFP)^26^. To improve on these previous studies, we capitalized on the fact that nanobodies (and modified versions thereof) can be efficiently expressed in *Escherichia coli*. We designed a recombinant protein where the well-characterized anti-GFP nanobody^27^ is fused, through a flexible peptide linker, to a SpyCatcher domain (**Fig. 2a**). This SpyCatcher domain is specifically recognized by a short SpyTag peptide, triggering the autocatalytic formation of a covalent isopeptide bond linking both of them with very high stability. Originally engineered from a bacterial adhesion protein^28^, the SpyTag/SpyCatcher protein conjugation system has been used for several applications^29^, but has not yet been applied to FET or OECT biosensors. The only related application of a nanobody-SpyTag fusion was a passive, impedance-based sensor for the detection of microalgae^30^ which relied on the random chemical immobilization of separated SpyCatcher proteins as a capture reagent.

For our platform, we wanted to avoid steps where the proteins (nanobody or SpyCatcher) are chemically modified or immobilized in a way that could partially impair their function. Instead, we opted for the immobilization of a chemically modified SpyTag, obtained through regular commercial peptide synthesis, on a 1,6-hexanedithiol (HDT) self-assembling monolayer, thus forming a combined Chem-SAM on top of the gold gate electrode. The anti-GFP nanobody^27^-SpyCatcher fusion protein was genetically encoded, gene synthesized and produced in *E. coli*. The purified protein construct was then incubated under physiological conditions with the Chem-SAM, thus completing the sensing surface with a self-assembled Bio-SAM. This strategy leads to a defined biofunctionalization layer in a precisely controlled molecular configuration and orientation, which can be modelled at atomic resolution (**Fig. 2b**). A flexible 8-amino acid glycine-serine linker separates nanobody and SpyCatcher domains dynamically and sterically. Taking into account unstructured residues contributed by the nanobody and SpyCatcher domains, the 4 nm long nanobody domain is separated from the SpyCatcher adapter by about 30 amino acids, forming a ∼12 nm long flexible molecular leash (**Fig. 2a,b**). Additional flexibility is introduced by short glycine-serine and alkane linkers that separate the chemically synthesized SpyTag sequence from the underlying HDT, thus also giving the SpyCatcher domain some freedom to move and rearrange itself above the Chem-SAM.

The three immobilization steps (HDT, SpyTag, nanobody-SpyCatcher) were monitored through X-ray photoelectron spectroscopy (XPS), quartz crystal microbalance with dissipation (QCM-D), cyclic voltammetry (CV), and electrochemical impedance spectroscopy (EIS). Upon functionalization with the HDT layer, the gold electrode displayed characteristic thiol-gold (S-Au) and free thiol (-SH) peaks in its high-resolution S 2p XPS spectrum^31,32^, indicating the upright orientation of HDT linked to gold through only one of the two -SH moieties (with the other terminal pointing away from the surface) (**Fig. 2c)**. As we successively introduced SpyTag peptide and then the nanobody fusion protein on top of the Chem-SAM, a new peak appeared at about 163 eV, which we attributed to the S-C bond from protein methionine residues (**Fig. 2c)**^33^. High resolution C 1s and N 1s XPS spectra further demonstrated the Bio-SAM formation by revealing C-O, C-N, C-OOR bonds, and nitrogen groups on the surface (**Fig. S1)**. Formation of all layers was further corroborated by EIS and CV measurements, which showed a decrease in the electrochemical capacitance of the gold electrode and an increase in its charge transfer resistance with the addition of (insulating) Chem-SAM and Bio-SAM (**Fig. S2**).

To assess the packing density of biomolecules on the surface, we monitored the immobilization of, first, the maleimide-modified SpyTag peptide and, second, the nanobody-SpyCatcher fusion protein on the Chem-SAM by QCM-D. We quantified the mass gained from the two conjugates to be 114 and 406 ng cm^-2^, respectively (**Fig. 2d and Fig. S3**). Based on a molecular weight of 1.76 kDa and 28.4 kDa for the SpyTag peptide and nanobody-SpyCatcher protein, respectively, 39 × 10^12^ SpyTag peptides and 8.6 × 10^12^ nanobody-SpyCatcher molecules were coupled per cm^2^. This density translates to only 3.4 nm mean inter-particle distance between the SpyCatcher domains at the base of the Bio-SAM. Given the size of this domain (about 4 × 2.5 nm), we are thus approaching the maximum coupling density that is physically feasible. The formation of this exceptionally high-density biorecognition layer is likely further facilitated by the compact nanobody, which has similar dimensions to the SpyCatcher domain but is given additional freedom to pack and reposition itself through the flexible inter-domain linker (**Fig. 2b**).

## Optimization of protein detection

We used the detection of GFP with the anti-GFP nanobody-conjugated gate electrode as a model to optimize multiple parameters of the sensor design. Initial optimization rounds addressed unspecific binding which was eliminated by adding bovine serum albumin (BSA) and a mild detergent (Tween-20) to both the nanobody-SpyCatcher protein immobilization and the target-binding solutions. The co-addition of BSA with SpyCatcher protein outperformed other blocking strategies. Comparing QCM-D binding traces with (**Fig. 2d**) and without BSA (**Fig. S3**), we suggest that BSA may be primarily capturing contaminating proteins in solution rather than binding to and blocking the surface in a classical sense. We next optimized the OECT geometry through systematic variation of gate and channel sizes. The highest sensitivity towards GFP was attained with an OECT having a gate electrode area of 0.64 mm^2^, and a channel width of 100 µm and length of 10 µm (**Fig. S4)**.

The optimized PEDOT:PSS OECTs were operated in PBS under ambient conditions by varying the gate voltage (*V*_G_) between -0.6 V and 0.6 V while the drain voltage (*V*_D_) was swept from 0 to -0.6 V. We assessed the sensing signal by monitoring the drain current (*I*_D_) as a function of the *V*_G_ at a fixed *V*_D_ = -0.6 V. The reference (blank) response of the sensor was acquired by exposing the nanobody-functionalized gate electrode to the binding buffer without GFP (**Fig. 3b**). The same electrode was then incubated for ten minutes with a 5 µL drop of binding buffer containing GFP, rinsed twice in PBS, and then stacked vertically on top of the channel separated by PBS (phosphate-buffered saline, pH 7.4, ionic strength 0.162 M) to complete the OECT biosensor (**Fig. 1a**). After gate exposure to GFP, *I*_D_ underwent a notable reduction at all gate voltages applied. As the GFP concentration increased, this sensor response became more evident (**Fig. 3b**). Similarly, the transconductance of the devices decreased as GFP interacted with the gate electrode (**Fig. S5a**). In contrast, the device characteristics did not change if the gate electrode was incubated in solutions containing equivalent molar concentrations of non-target molecules such as an FRB-mCherry fusion (**Fig. 3c, Fig. S5b**) and lysozyme (**Fig. S6**). Lysozyme is abundant in saliva which we envisioned as a potential biological medium for our SARS-CoV-2 application. mCherry is a red fluorescent protein with low sequence homology but very high structural similarity to GFP and thus challenged sensor specificity.

**Fig. 1.**
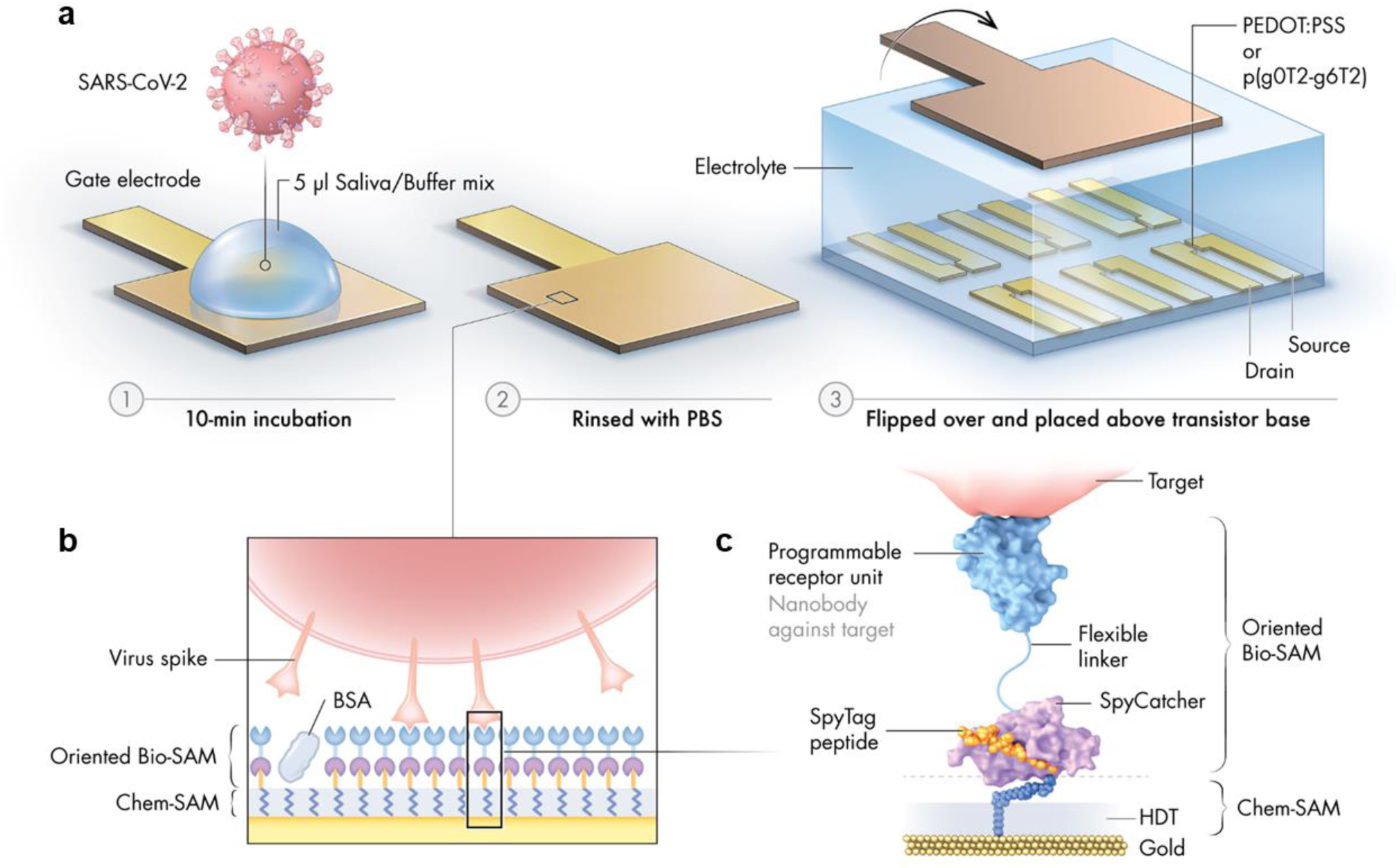
Schematic illustration of the nanobody-functionalized OECT sensor. **a)** Operation: The gate electrode is exposed to a mix of sample (such as saliva) and binding buffer (1), rinsed with PBS (2), and mounted on top of the OECT channel for signal acquisition (3). **b**) Gate functionalization layers. Chemical and biological monolayers (Chem-SAM and Bio-SAM) are self-assembled on the gate electrode surface. **c)** Molecular architecture. A synthetic SpyTag peptide is chemically coupled to the 1,6-hexanedithiol (HDT) monolayer to form a Chem-SAM. The nanobody-SpyCatcher fusion protein then attaches itself to this chemical layer through the autocatalytic formation of a covalent SpyCatcher-SpyTag bond, forming the Bio-SAM. The nanobody domain defines sensor specificity and is interchangeable. Bovine serum albumin (BSA) is physisorbed on the sensor surface during the final step of functionalization to prevent non-specific binding.

**Fig. 2.**
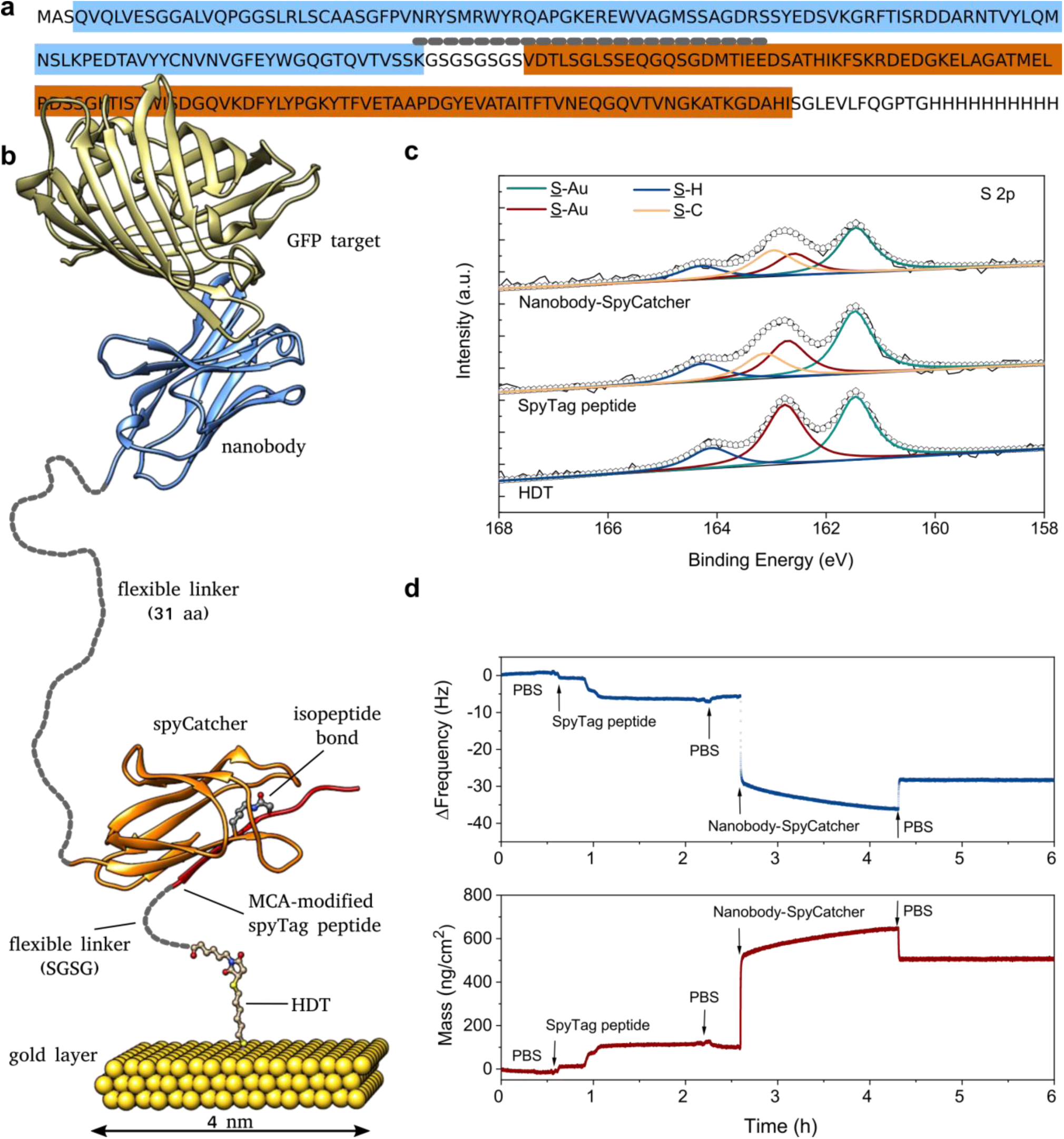
Design and characterization of the biofunctionalized gold electrode. **a)** Annotated sequence of the recombinant fusion protein combining a nanobody recognition module (blue) with the SpyCatcher domain (brown) with an intervening flexible linker (short-dashed line). **b)** Structural model of the complete biorecognition layer assembled from HDT and SpyTag (Chem-SAM) and nanobody-SpyCatcher fusion protein (Bio-SAM) (based on PDB structures 4MLI and 3OBO). Unstructured, flexible linker regions are indicated by short-dashed lines drawn to scale. **c)** High resolution S 2p XPS spectra of the gold electrode recorded after immobilization of HDT, SpyTag peptide, and the nanobody-SpyCatcher protein using final conditions with BSA added to the immobilization. **d)** QCM-D profile measuring the coupling of SpyTag peptide and nanobody-SpyCatcher fusion protein (without BSA) in real-time, starting from an HDT-coated gold electrode.

**Fig. 3.**
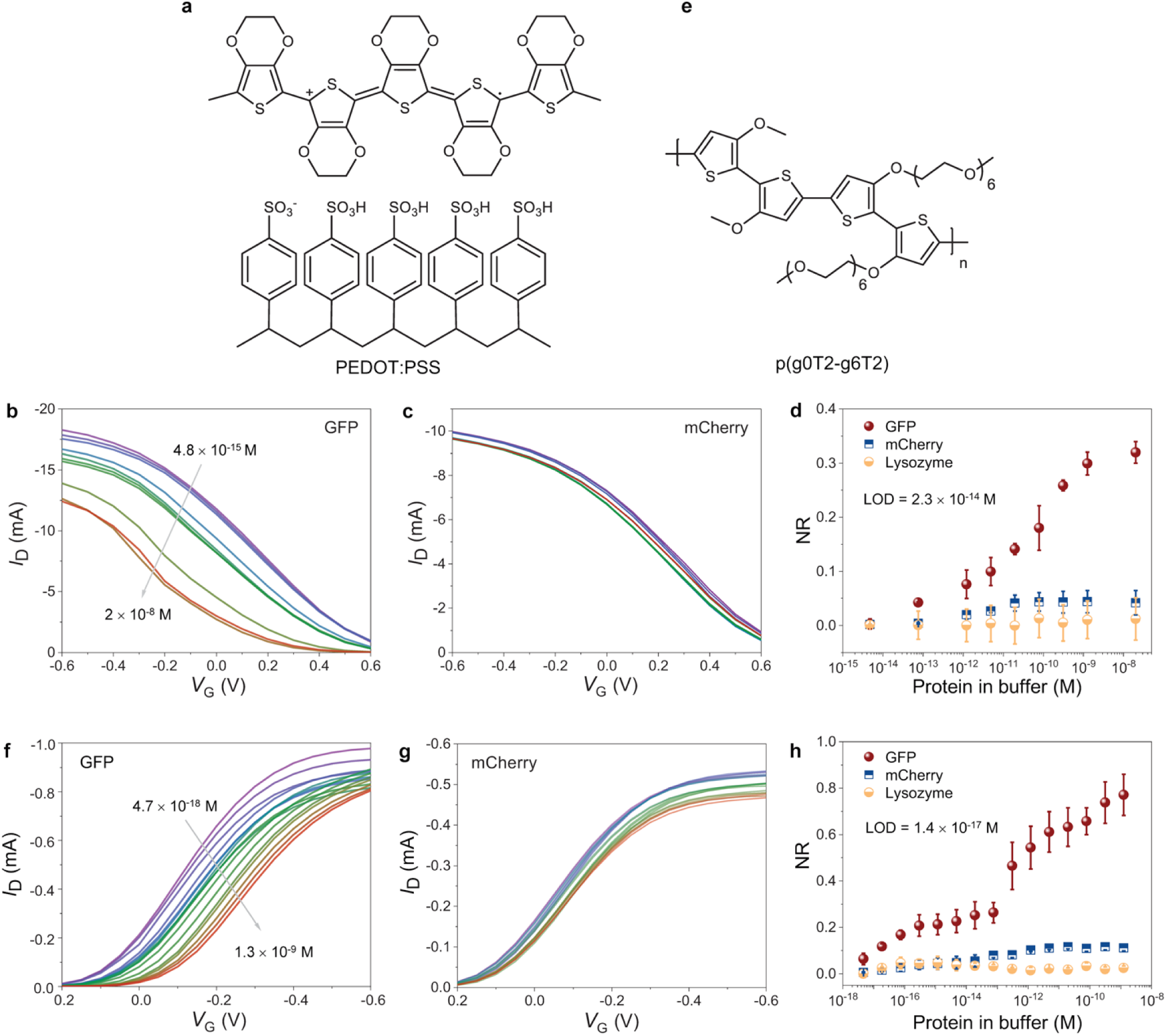
Performance of GFP nanobody-functionalized OECT biosensors. Chemical structure of **a)** PEDOT:PSS and **e)** p(g0T2-g6T2). The steady-state characteristics of OECTs gated with a GFP-functionalized gold electrode exposed to **b), f)** GFP, and **c), g)** mCherry. **d), h)** The normalized response (NR) of the sensors to GFP, mCherry, and lysozyme. In **b-d**, the channel is PEDOT:PSS, **f-h)** the channel is p(g0T2-g6T2). Protein concentrations in the buffer range from 5 fM to 20 nM. In b and f, the decrease in *I*_D_ (and the increase in protein content in the buffer) is indicated by the direction of the arrow. Error bars represent the standard deviation calculated using at least 3 gate electrodes.

The trend in *I*_D_ and *g*_m_ can be clearly attributed to the specific binding of GFP to the nanobody-functionalized gate electrode. Fluorescence images confirmed the specific capture of GFP, but not mCherry (**Fig. S7**). The GFP-nanobody interaction increases the impedance at the gate electrode/electrolyte interface (**Fig. S8**), resulting in a voltage drop therein and a less efficient capacitive coupling between the gate and the channel. Hence, we measure a reduction in the current and its modulation as GFP-binding weakens the electrical driving force acting on the cations. Torsi and co-workers observed similar changes in PEDOT:PSS OECT characteristics, where the *I*_D_ decreased upon a protein binding event at the functionalized gate electrode^20^. We note that the decrease in *I*_D_ is not due to a degradation of the channel over consecutive I-V cycles (**Fig. S9**). Neither did we observe a change in gate current (*I*_G_) upon GFP binding, and the value of *I*_G_ was six orders of magnitude lower than the ON current of the OECT (**Fig. S10**). To quantify the sensor response and minimize device-to-device variations, we calculated the normalized response (NR) of the OECT by normalizing the target protein-induced change in *I*_D_ at a single *V*_D_ and *V*_G_, to its value previously measured after exposure to the blank solution. The NR *vs*. GFP calibration curve revealed that the nanobody-functionalized OECT detects GFP with a lowest limit of detection (LOD) of 23 fM **(Fig. 3d)**. The NR was less than 5% even for the highest concentrations of negative controls (mCherry and lysosome) while it exceeded 30% for higher GFP concentrations. Overall, PEDOT:PSS-based OECT biosensors were sensitive to the presence of 6 × 10^4^ GFP molecules in 5 µL of buffer and responded within a dynamic range of six orders of magnitude. This sensitivity is on-par with commercial colorimetric ELISA assays which, however, take hours to complete, require larger sample volumes and have a far narrower dynamic range^34^.

## Sensing with accumulation mode OECTs

As PEDOT:PSS is intrinsically doped, our biosensors operate in depletion mode, that is, *I*_D_ decreases with an increase of *V*_G_. High OFF currents and the *V*_G_ applied to keep the device in its off-state increase power consumption and may diminish material stability during long term use. Recent studies expanded the available OECT channel materials through the chemical design of undoped, conjugated backbones which were functionalized with hydrophilic side chains that facilitate ion injection and transport in the film^16,35^. OECTs comprising these materials operate in accumulation mode, that is, the channel is OFF and generates a current upon application of a small gate voltage^17^. This operation mode allows for low-power electronics applications, improves device stability, and is more compatible with integrated circuit designs that conventionally involve accumulation mode transistors. Nevertheless, accumulation mode OECTs for biochemical sensing have not yet been reported. We, therefore, replaced PEDOT:PSS with a recently designed conjugated polymer, p(g0T2-g6T2)^36^ (**Fig. 3e**). p(g0T2-g6T2) is a mixed (ionic and electronic) conductor. A negative *V*_G_ pushes anions into the film that compensate for the holes injected from the metal contacts, turning the device ON (**Fig. S11**). The maximum *g*_m_ occurs at a *V*_G_ of ca. -0.3 V and the transconductance efficiency (*g*_m_ obtained per unit current), is very high at low *V*_G_ due to the exponential *I*_D_ vs. *V*_G_ characteristics. This behavior is similar to the subthreshold region of operation in traditional inorganic metal oxide semiconductor FETs^37^. The material also shows exceptional operational and environmental stability (**Fig. S12**), thus overcoming the limitations of the currently available organic mixed (semi)conductors^36^.

**Fig. 3f** shows the typical transfer characteristics of p(g0T2-g6T2) OECTs gated with the nanobody-functionalized gold electrodes incubated in solutions with varying GFP concentrations. As GFP bound to the gate, *I*_D_ decreased for all gate voltages. This decrease was accompanied by a significant shift in the threshold voltage (*V*_th_) towards more negative values (**Fig. S13**). Similar to PEDOT:PSS sensors, p(g0T2-g6T2) OECTs showed no significant response to mCherry (**Fig. 3g**) and lysozyme (**Fig. S14**) (at closer inspection, our data suggest a minor cross-reaction of the anti-GFP nanobody with the structurally similar mCherry). The sensor responded to a GFP concentration as low as 4.7 aM (NR = 7%) (**Fig. 3h**) with a dynamic range spanning 10 orders of magnitude. We calculated the lowest limit of detection (LOD) for these devices to be 14 aM, corresponding to 42 molecules in the 5 μL sample. Compared to the otherwise identical PEDOT:PSS-based sensor, the accumulation mode OECT thus showed a 1000-fold improved sensitivity (aM compared to fM) and yet larger dynamic range (10 orders of magnitude, from aM to nM, compared to 6 orders of magnitude). Moreover, we noted a much lower operating voltage. The target response was maximized (and off-target response minimized) at *V*_G_ = *V*_D_ *=* -0.1 V compared to *V*_G_ = *V*_D_ *=* |0.6*V*| for the PEDOT:PSS OECT (**Fig. S15**). Such characteristics would, in principle, allow for nanobody-OECT reader devices with very compact built and long battery life.

From a biochemical point of view, the observation of binding at such low concentrations was initially surprising. The 14 aM detection limit lies 30 million-fold below the consensus equilibrium dissociation constant (K_D_ ∼0.5 nM) reported for the interaction between nanobody and GFP^38^. However, given the density of nanobody molecules (8.6 × 10^12^ / cm^2^, see above) and assuming the domain is diffusing within a 20 nm layer above the sensor surface (**Fig 2b**) we arrive at a local nanobody concentration of about 700 µM. Any target molecule diffusing into this capture layer will therefore necessarily bind. Once bound, the interaction*’*s low k_off_ rate^38^ (∼1.5 × 10^−4^ s^-1^ corresponding to 80 min half-life) will trap the target molecule throughout all washing steps. In fact, our surface plasmon resonance (SPR) measurements indicate an even lower, essentially unmeasurable, off rate for this complex with our own protein and buffer conditions (**Fig. S16**). The high-density nanobody-functionalized gate electrode thus operates as a kinetic trap for target molecules. Sensitivity is likely not primarily dictated by nanobody:target affinity (K_D_) but by target diffusion and the kinetic stability of the complex (k_off_). We were thus hopeful to reproduce sensor performance with other nanobodies.

## Detection of SARS-CoV-2 and MERS-CoV antigens

The VHH72 nanobody was originally raised against SARS-CoV receptor binding domain (RBD). Subsequently, VHH72 was shown to also bind SARS-CoV-2, at a slightly reduced affinity, and it became the first publicly available nanobody sequence against this new target^39^. Our VHH72 -spyCatcher fusion construct expressed well in *E. coli* and could be purified to a high yield (54 mg/L of culture) as a monomeric protein. Although SARS-CoV-2 was our primary diagnostic target, we also designed and expressed constructs for the detection of MERS-CoV based on the previously reported nanobodies VHH83^40^, VHH04^40^, and VHH55^39^. The VHH04-spyCatcher fusion yielded high-quality monomeric protein and was advanced to OECT experiments. Both VHH72 and VHH04 bind to the RBD of the homotrimeric (SARS or MERS, respectively) Coronavirus Spike (S) protein^41^ which protrudes from the virus surface in multiple (about 100) copies. The RBD is a small (27 kD) protein domain within the larger S1 subunit (76 kD) and is directly responsible for the recognition and binding of the specific host cell receptor. The detection of RBD would therefore be a good indicator for the presence of infectious viral particles. SPR experiments confirmed the binding of both nanobody fusion proteins with their respective, recombinantly expressed, SARS-CoV, SARS-CoV-2 or MERS-CoV target proteins (**Fig. S17**).

As expected, VHH04 showed the highest affinity (K_D_ =0.1 nM) and slowest k_off_ (2×10^−4^ s^-1^) for its target MERS-CoV S1. VHH72 bound its primary target SARS-CoV(1) RBD with lower affinity (K_D_ =7 nM), owing to a faster dissociation (3×10^−3^ s^-1^). The interaction with the closely related RBD of SARS-CoV-2 showed a mixed signal, where a smaller share of fast-on, fast-off binding events was distorting the main slow-on, slow-off binding regime (K_D_ =23 nM, k_off_ =3×10^−4^ s^-1^). Such a signal was also reported for the interaction with a more recently developed nanobody^42^ and may indicate an intrinsic heterogeneity of this RBD.

Nevertheless, SARS-CoV nanobody-functionalized OECTs showed an excellent response to the SARS-CoV-2 RBD and S1 subunit, regardless of the channel material used (**Fig. 4a and b**). Incubation of the same gate electrodes with various concentrations of GFP did not evoke any current response, demonstrating the high specificity of the VHH72-functionalized gate for the viral proteins. The device also responded to the original target of VHH72, the SARS-CoV-1 RBD (**Fig. S18)**. In direct comparison, the higher affinity of VHH72 for SARS-CoV-1 RBD compared to SARS-CoV-2 RBD did only translate into a small signal increase. By contrast, the larger SARS-CoV-2 S1 spike protein (comprising the same RBD) generated much larger current changes than the isolated RBD. This increase in NR may stem from higher affinity or size or both. We assume that the S1 subunit will, at least partially, form trimers in solution (all concentrations given in this study refer to the monomer) and thus benefit from avidity effects with substantially increased affinity (and prolonged residence) on the gate surface. Compared to RBD alone, both the S1 monomer or trimer would also cover a larger area on the functionalized surface than the isolated RBD. Akin to our GFP sensors, p(g0T2-g6T2) OECTs showed higher sensitivities than PEDOT:PSS devices. p(g0T2-g6T2) OECT detected SARS-CoV-2 S1 at 4.7 aM with a 30% change in the NR (*σ*_*SD*_ = 7% at most), translating to a nominal LOD of 18 zM. In fact, a single molecule in our 5 µL measurement volume corresponds to a concentration of 3.3 aM which appeared to be easily detected.

**Fig. 4.**
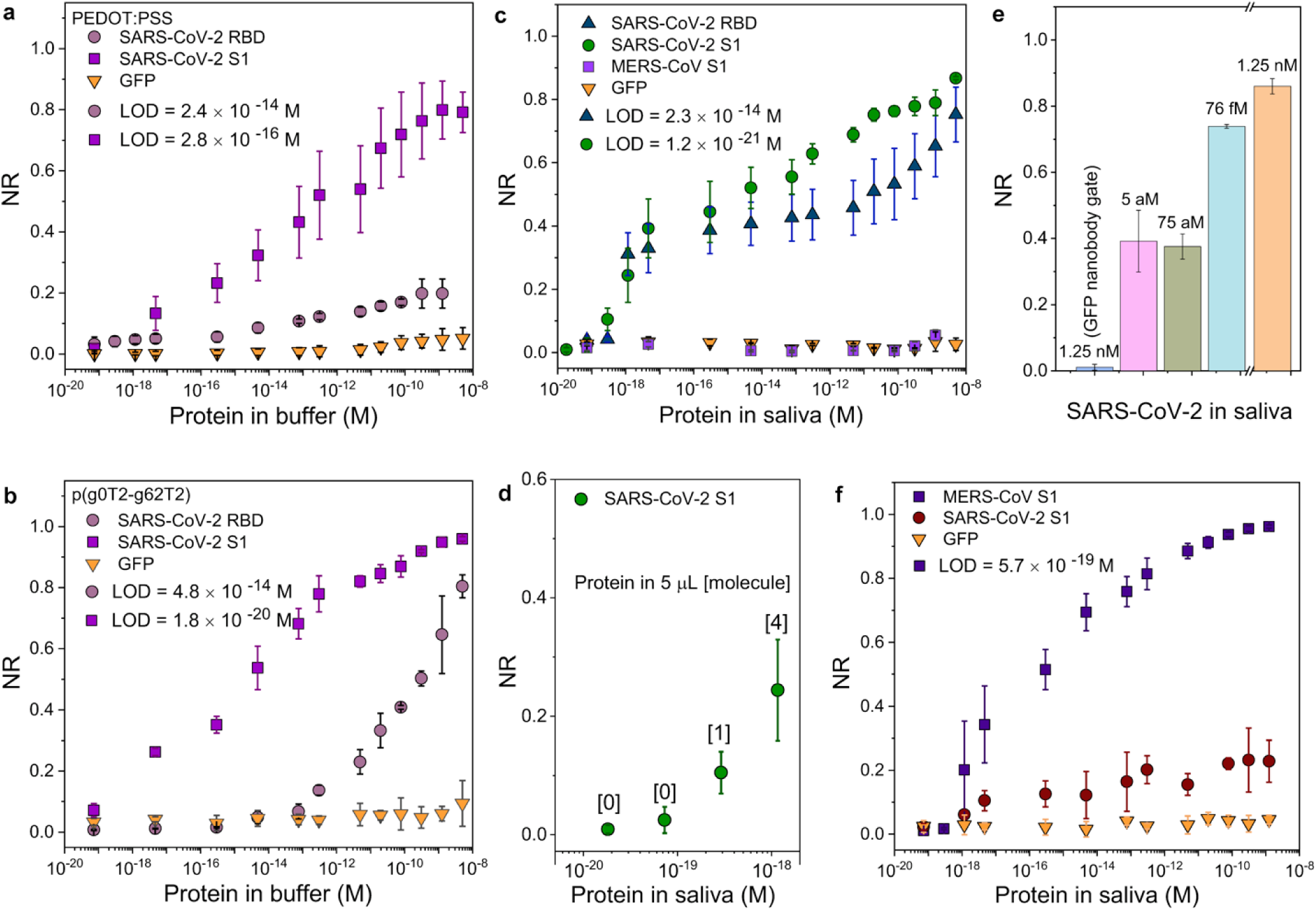
Performance of the SARS-CoV-1 or MERS nanobody-functionalized OECTs in detecting SARS-CoV-2 or MERS-CoV. The normalized response (NR) of SARS-CoV nanobody-functionalized OECTs to SARS-CoV-2 RBD, S1, and GFP for channels comprising **a)** PEDOT:PSS and **b)** p(g0T2-g6T2). **c)** The response of SARS-CoV nanobody-functionalized OECTs comprising p(g0T2-g6T2) channel to SARS-CoV-2 proteins, GFP and MERS-CoV S1 spiked in human saliva. **d)** The NR of SARS-CoV-2 sensors at single molecule detection level. **e)** The NR of SARS-CoV-2 sensors to randomly-selected saliva samples containing various amounts of SARS-CoV-2 S1. One of the samples (1.25 nM of target protein) was measured with GFP nanobody gate. The blank measurements were obtained by incubating the gate electrodes in PBS.**f)** The response of MERS nanobody-functionalized OECTs comprising p(g0T2-g6T2) channel to MERS S1, SARS-CoV-2 S1 and GFP spiked into human saliva. In all measurements, error bars represent the standard deviation calculated from measurements with at least 3 gate electrodes.

Having confirmed single-molecule sensitivity and high specificity of SARS-CoV nanobody OECTS when target proteins were captured in the binding buffer, we turned our attention to sensor performance in saliva. Saliva samples are reported to exhibit comparable or higher viral loads than nasal swaps^43,44^. While this may not lead to perfect sensitivities under real-world sampling conditions^45^, saliva is clearly the better medium for point-of-care or field applications. We, therefore, challenged SARS-CoV nanobody-functionalized OECTs comprising p(g0T2-g6T2) channels with human saliva samples into which we spiked predetermined amounts of target and non-target proteins. The NR increased with the concentration of SARS-CoV-2 proteins in saliva and showed only negligible change when the same gate electrode was exposed to GFP (**Fig. 4c)**. As before, sensor response already picked up at the single-molecule concentration threshold (**Fig. 4d**). The sensitivity for SARS-CoV-2 S1 in saliva (LOD = 1.2 × 10^−21^ M) was comparable to that in buffer (LOD = 1.8 × 10^−20^ M) indicating that the complex composition of saliva did not hamper the association between the nanobody and its target. The OECT could also detect SARS-CoV-2 S1 mixed into human serum (**Fig. S19**), indicating its potential use for a wide range of other diagnostic applications.

So far, we have characterized sensor responses through the repeated exposure of one gate electrode to increasing target concentrations (all results are averages over several electrodes gating several channels each). In order to better emulate real-world screening conditions, we directly exposed individual gate electrodes to a single random concentration of SARS-CoV-2 S1 in saliva (**Fig. 4e**). The NR values varied accordingly and corresponded to the values obtained in the regular dose curves. By contrast, even high concentrations of SARS-CoV-2 S1 did not evoke any response on a gate functionalized with GFP nanobody.

MERS-CoV is the only close relative of SARS-CoV-2 in active circulation^46^. The MERS-CoV nanobody-functionalized gate electrode detected single MERS-CoV S1 protein molecules in saliva, with a nominal detection range from 100 zM to 1 nM (**Fig. 4f**). A weak cross-reaction of VHH04 with SARS-CoV-2 target protein could already be anticipated from SPR measurements (**Fig. S17**). By contrast, despite the high structural similarity between the two proteins, the SARS-CoV sensor did not respond to MERS-CoV S1 (**Fig. 4c**). Remarkably, the MERS-CoV sensor again achieved single-molecule sensitivity and 10 orders of magnitude dynamic range in complex biological media without prior optimization to this particular target (see **Table S1** for the LODs of all the sensors used in this study). Our findings thus demonstrate the sensitivity and selectivity of nanobody-OECTs in complex biological media and at physiologically and clinically relevant protein concentrations. We expect that the nanobody-OECT platform can be rapidly adapted to detect any protein antigen for which nanobodies are available.

## Detection of SARS-CoV-2 in Clinical Samples

We validated our sensors using human nasopharyngeal swab and saliva samples collected in a hospital setting. Swabs were received from clinicians in a commercial universal transport medium (UTM) and analyzed after two days. A calibration curve with SARS-CoV-2 S1 protein confirmed sensor operation in this UTM, albeit at lowered sensitivity (LOD for SARS-CoV-2 S1 in UTM was 1.9 × 10^−14^ M) (**Fig. S20**). Saliva was collected from healthy volunteers and from COVID-19 in-patients (at different stages of disease) without instructions or protocols from our side. Control measurements were performed with the same samples using GFP-nanobody gate electrodes and viral status was verified independently by RT-PCR. In all but one case, sensor readings agreed with RT-PCR on the presence or absence of SARS-CoV-2 in the sample (**Fig. 5a-b, Fig. S21**). In the remaining case (saliva sample #10), our sensor gave a clear positive diagnosis where the RT-PCR result was negative. The saliva sample in question had been collected from a hospitalized COVID-19 patient, but was (like many other samples) partially dried-out by the time of measurement. Low saliva sample quality likely impacted both sensor and RT-PCR experiments. By contrast, four nasal swab samples that had previously been tested as positive by RT-PCR were also confirmed to be positive by the OECT. Notably, the four positive nasal swabs had low to very low viral loads (RT-PCR cycle threshold, CT, 33.4 to 39, close to the CT=40 detection limit). For a direct comparison of detection limits, we prepared a 10-fold dilution series in UTM of one patient sample. We mixed each dilution 1:1 with binding buffer and subjected it side-by-side to sensor measurements as well as RNA extraction & RT-PCR (**Fig. 5c**). The sensor*’*s above-noise response appeared to outperform RT-PCR by one order of magnitude. The experience with real-world samples thus suggests sensor performance on-par with RT-PCR but also points to some areas that need to be optimized along the path from a lab-based prototype to an actual diagnostic device.

**Fig. 5.**
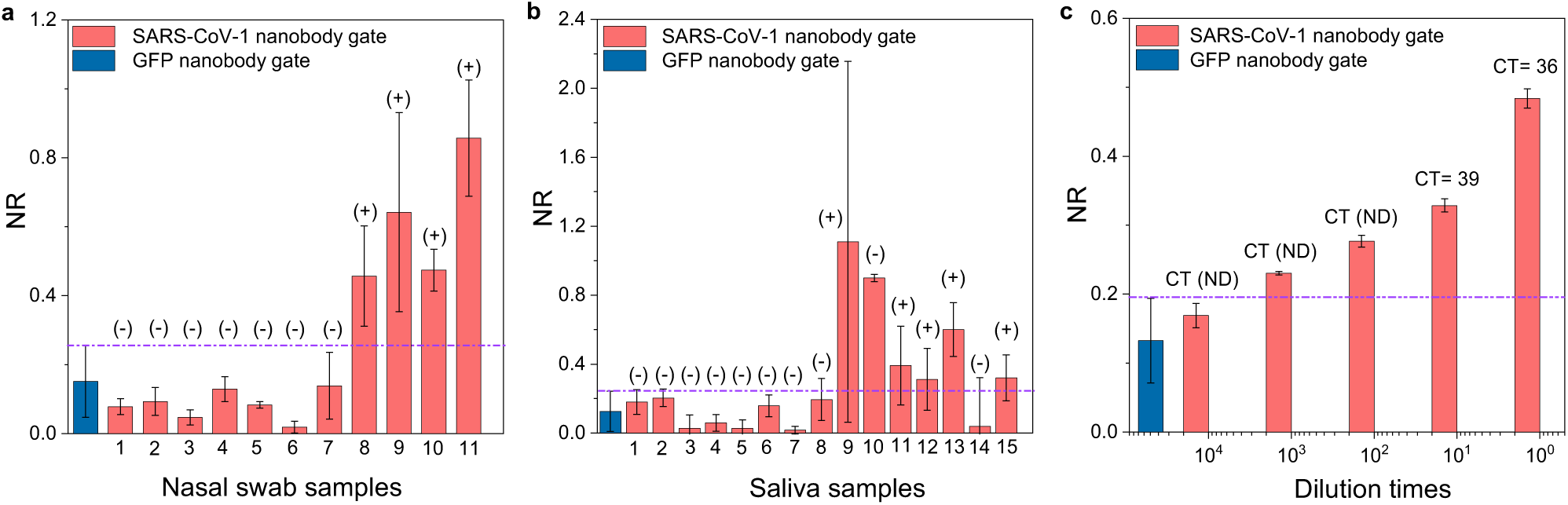
SARS-CoV-2 detection in clinical samples with OECT biosensors. Normalized sensor response (NR) for **a)** nasopharyngeal swab samples (n=11) from healthy volunteers (1-7) and from walk-in patients tested SARS-CoV-2 positive by RT-PCR (8-11); **b)** saliva samples (n=15) taken from healthy volunteers (1-7) and from hospitalized COVID-19 diagnosed patients (8-15). RT-PCR results for the same samples are indicated (+, positive) and (-, negative). All RT-PCR results were CT > 30, indicating very low viral loads. The control experiments were performed by testing the same specimens with GFP gate electrodes and the error bars represent the average results from at least 12 electrodes. The threshold line indicates mean current + standard deviations of signals collected from these control sensors. If the current change for any sample was higher than the threshold, the sample was considered as SARS-CoV-2 positive. **c)** Sensor results from a 10-fold dilution series of nasal swab sample #8. RT-PCR results (Cycle threshold, CT) after RNA extraction from the same sample are given above each bar. ND = not detected. Error bars for the SARS-CoV-1 gates represent standard deviations determined from measurements with 2 gate electrodes and 3 channels for each sample.

## Conclusion

We here introduced a label-free electrochemical immunosensing platform with single protein molecule sensitivity and large dynamic range. The nanobody-OECT platform detects specific protein molecules from unprocessed samples in ambient conditions after only 10 minutes of incubation with the disposable biofunctionalized gate electrode. The sensor can be rapidly adapted to any target for which nanobodies exist or can be raised. After optimizing the sensor for GFP detection, the simple exchange of nanobodies gave attomolar sensitivities for SARS-CoV-2 and MERS spike proteins in saliva or buffers without any further modification to the sensor or protocol. Preliminary tests with samples from COVID-19 patients and healthy volunteers demonstrate a sensor accuracy and sensitivity on par with RT-PCR. For COVID-19, the detection of intact spike protein may, in fact, be a better indicator of infectious virus than RT-PCR, as the latter is also detecting membrane-associated viral RNA fragments long after the infection has been cleared^47^.

We attribute the performance of our sensor to several synergistic design choices: The OECT architecture outperforms other transistor technologies in terms of signal amplification and responds to changes far from the sensor surface despite its simplicity and miniaturized geometry. These characteristics allowed us to tether biorecognition domains through long flexible linkers. A novel bioconjugation strategy, based on the autocatalytic assembly of a protein adapter with a chemically synthesized peptide, allowed for the oriented coupling of chemically unmodified recognition proteins at an exceptionally high density limited only by the size of the proteins used. The choice of nanobodies over classic antibodies (or antibody fragments) further improved the density and robustness of the biorecognition layer. The resulting three-dimensional and flexible capture layer displayed a kinetically controlled target binding regime that is not limited by dissociation constants. Moreover, a new solution-processable organic semiconductor enabled the sensor to operate in accumulation mode which further improved sensitivity and stability by providing large changes in output current for operation at very low biasing conditions.

Several additional features prepare this sensor architecture for POC applications. The transistor base capable of recording from multiple micron-scale channels is reusable, allowing a stable electronic base to be combined with disposable biofunctionalized gate electrodes. Power consumption of the sensor is minimal and compatible with a hand-held battery-driven reader. Test results are currently obtained in less than 15 minutes, and we expect that changes in our 10-min incubation protocol can further accelerate the procedure. Real-time measurements are technically feasible when the platform is integrated with microfluidics. The functionalization is simple and modular, the device is easy-to-fabricate and obviates complex operation steps. The speed, performance and versatility of our nanobody functionalized OECT, and its compatibility with many sample types, suggest that this platform can complement or replace a wide range of clinical and non-clinical diagnostic assays.

## Methods

### Materials

Sodium chloride, Tween-20, glycerol, HEPES, bovine serum albumin (BSA), (3-glycidyloxypropyl) trimethoxysilane (GOPS), sodium dodecylbenzenesulfonate (DBSA), ethylene glycol (EG), 1,6-hexanedithiol (HDT), 1,3-propanedithiol (PDT), 3-mercaptopropionic acid (MPA), human serum, and PBS (pH 7.4) were purchased from Sigma-Aldrich and used as received. Poly(3,4-ethylenedithiophene) doped with poly(styrene sulfonate), PEDOT:PSS, (PH1000) was received from Heraeus. All aqueous solutions were prepared with ultrapure water (Millipore Milli-Q). p(g0T2-g6T2) was synthesized according to a procedure reported previously^36^. Protein purification materials: Agar, LB broth, 2xYT Broth, Kanamycin, Glucose, Isopropyl β-D-1-thiogalactopyranoside (IPTG), BugBuster (Novagen), cOmplete Protease Inhibitor mix (Sigma), Benzonase (Novagen), Egg-Lysozyme (Fluka), Tris(2 carboxyethyl)phosphine (TCEP), Tris(hydroxymethyl)aminomethane hydrochloride (Tris-HCl), Imidazole, Glycerol, Dithiothreitol (DTT), Ethylenediaminetetraacetic acid (EDTA), D-Desthiobiotin, 10K Amicon ultra spin concentrators (Milipore). Purification columns and SPR materials were purchased from GE Healthcare: HisTrap-HP 5 ml, StrepTrap-HP 5 ml, Superdex75 16/600, Biacore NTA SPR sensor chips (BR100034). MCA-SpyTag peptide (Genscript, peptide synthesis). Viral target proteins were purchased from Sino Biological: SARS-CoV RBD (40150-V08B2), SARS-CoV-2 RBD (40592-V08H), SARS-CoV-2 S1 (40591-V08B1), MERS-CoV S1 (40069-V08H). Universal Transport Medium (UTM) Kit was purchased from Noble Biosciences, Inc.

### Fabrication of the organic electrochemical transistor (OECT) and the gate electrode

OECTs were microfabricated on glass substrates based on established protocols using standard photolithography and Parylene-C peel-off techniques^48,49^. The process starts with the first layer of photoresist (AZ5214) spin coated and exposed to UV light using contact aligner to create Au electrodes and interconnection pads. The photoresist patterns were generated with AZ 726 MIF developer, followed with metal sputtering of 10 nm of Cr and 100 nm of Au and a standard lift-off process using hot DMSO. We next coated the second layer of photoresist AZ9260 on the substrates and developed using AZ developer. A parylene-C layer was deposited to insulate the gold interconnects. The OECT channel was patterned by reactive ion etching (RIE) and using a second layer of a Parylene C which was peeled-off to yield a length of 10 µm in and width of 100 µm. The aqueous dispersion of poly(3,4-ethylenedioxythiophene) doped with poly(styrene sulfonate) (PEDOT:PSS) containing EG (5 vol%), DBSA (0.25 vol%), and GOPS (1 wt%) was sonicated for 30 min and then spin coated (3000 rpm, 45s) on the substrates leading to a film thickness of about 160 nm. The PEDOT:PSS OECTs were annealed at 140 °C for 1 h to activate GOPS and avoid dissolution of the polymer film in aqueous medium. P(g0T2-g6T2) films were spin coated (800 rpm, 45 s) from a chloroform solution (5 g/L) on the substrates to yield a film thickness of about 70 nm in the channel. All devices were rinsed with DI water before use.

For the fabrication of the gate electrode, we used 175 µm-thick Kapton (polyimide) substrates. The Kapton was first cut with a CO_2_ laser (Universal Laser Systems – PLS6.75) into a circular geometry that defined the sensor active area (the final diameter used in sensors is 0.8 µm). We then sonicated the substrates, first in isopropyl alcohol and then in deionized (DI) water for 30 min each. We sputtered 10 µm of chromium or titanium and 100 µm of Au on these cleaned substrates. Before the functionalization, the electrodes were electrochemically cleaned in 10 mM sulfuric acid (H_2_ SO_4_) using cyclic voltammetry (CV). 10 CV cycles with a potential between -0.2 V to 1.2 V were applied at scan rate 100 mV s^-1^.

### Biofunctionalization of gate electrodes

The chemical SAM solution was prepared in absolute ethanol containing 1 mM of HDT, as described by others^50^. The Au electrodes were immersed in this solution for one hour^32^. Electrodes were rinsed with absolute ethanol and dried under a N_2_ stream. Electrodes were then incubated for one hour with the synthetic Maleimide-modified SpyTag peptide (0.1 mg/mL) in PBS, and washed in PBS. Electrodes were then incubated for one hour with Nanobody-SpyCatcher fusion protein (anti-GFP, anti-SARS-CoV-1, or anti-MERS-CoV) diluted to 50 µM in sensor binding buffer (100 mM HEPES pH 7.4, 150 mM NaCl, 0.05% v/v Tween-20, 0.02% w/v NaN_3_, 0.1% w/v BSA) and then rinsed with PBS. Nanobody-functionalized gate electrodes were stored for up to one week at 4 °C in the sensor binding buffer until use.

### X-ray photoelectron spectroscopy (XPS)

XPS analysis was performed using a KRATOS Axis SUPRA instrument equipped with a monochromatic Al Kα X-ray source (1468.6 eV). We operated the source at 75 W under UHV condition (∼10^−9^ mbar). The spectra were recorded in a hybrid mode using electrostatic and magnetic lenses and an aperture slot of 300 by 700 µm. The survey and high-resolution spectra were acquired at fixed analyzer pass energies of 80 eV and 20 eV, respectively. We mounted the samples in a floating mode to avoid differential charging. The spectra were then acquired under charge neutralization conditions. We calibrated the spectra to reference of C 1s at 284.8 eV and deconvoluted it using Gaussian and Lorentzian methods. The Tougaard method was used for the background subtraction.

### Quartz crystal microbalance with dissipation monitoring (QCM-D)

QCM-D measurements were conducted using a Q-sense analyzer (QE401, Biolin Scientific). The piezoelectrically active gold sensor (0.7854 cm^2^) was firstly modified with HDT SAM under the same reaction conditions and then mounted into the QCM -D setup. First, the QCM-D signals, including the change in frequency (Δ*f*) and dissipation (Δ*D*) were stabilized in PBS. Second, the peptide solution (0.1 mg/mL SpyTag peptide in PBS) was injected into the chamber with a flow rate of 100 µL/min controlled by a peristaltic pump. After ensuring that the sensor was fully covered with the solution, we stopped the pump for an hour and rinsed the sensor surface with PBS injected to the system for 15 min. The same procedure was employed to treat the surface with SpyCatcher-linked nanobody solution (50 µM in the binding buffer). To quantify the mass accumulating on the sensor (Δ*m*) during the functionalization steps, we used the Sauerbrey equation^51^:

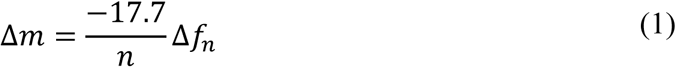

where *n* is the overtone number selected for the calculations and -17.7 is a constant calculated based on the resonant frequency, active area, density and shear modulus of the quartz crystal sensor. The mass of the binding molecules on the sensor surface was then estimated using their molecular weight.

### Electrochemical characterization

The electrochemical characteristics of the gold electrode were investigated before and after the formation of Chem-SAM and Bio-SAM using electrochemical impedance spectroscopy (EIS) and cyclic voltammetry (CV) performed in a three-electrode setup using a potentiostat (Autolab PGstat128N, MetroOhm). We used a platinum wire and an Ag/AgCl electrode as the counter electrode and reference electrodes, respectively, while the gold electrode was connected as the working electrode. All measurements were carried out in 5 mL of 10 mM PBS (pH 7.4) containing 10 mM of [Fe(CN)_6_]^3-/4-^. For CV measurements, the potential window was between -0.2 V and 0.5 V and the scan rate was 100 mV s^-1^. Electrochemical impedance spectra were recorded at a DC voltage of 0 V versus open circuit potential (V_oc_) and an AC modulation of 10 mV over a frequency range of 0.1-100000 Hz. For the analysis of GFP nanobody-functionalized electrodes, the electrodes were incubated with GFP solutions for 10 min and washed in 10 mM PBS before the measurements. The data was analyzed using Nova software.

### Proteins and Peptides

Nanobody-SpyCatcher fusion proteins were designed based on available structures (nanobody: PDB 4PFE^52^; SpyCatcher: PDB 4MLI^53^) with the nanobody placed at the N-terminal end of the fusion protein in order to orient the common VHH target-binding interface towards the bulk solution, away from the sensing surface. Protein sequences were reverse-translated and codon-optimized for expression in *E. coli* with an in-house Python script based on DNAChisel^54^. Plasmids for protein expression were gene synthesized by Twist Bioscience (U.S.A.) or BioBasic (Canada) in our customized expression vector pJE411c with kanamycin resistance and modified with a RBS insulator (BCD2) cassette^55^ for improved translation initiation. Plasmids were transformed into *E. coli* BL21 (DE3) and starter cultures were inoculated overnight from a single colony. 1L production cultures in 2xYT medium with 50 mg/L kanamycin and 1% glucose were inoculated 1:100, grown at 37°C and 250 rpm to OD_600_ 0.8, induced with 0.5 mM IPTG and incubated shaking for 18 h at 25°C. Cells were harvested by centrifugation for 10 min at 6,000 *g* at 4 °C, washed once with cold PBS, resuspended in lysis buffer [25 mM Tris-HCl (pH 7.4), 500 mM NaCl, 10 mM Imidazole, 10% glycerol, SigmaFAST protease inhibitor, 25 U/ml Benzonase HC (Milipore), 2 mM DTT], and homogenized with a cell disruptor (Constant Systems, UK). Earlier purifications of mCherry, GFPnanobody and msfGFP used different lysis methods (chemical lysis with BugBuster (Novagen) for mCherry, sonication for the other two). Lysates were cleared by centrifugation at 87,000 *g* for 45 min, the supernatant filtered through Miracloth tissue (Milipore) and subjected to affinity chromatography on an Äkta FPLC (GE Healthcare) using either StrepTrap HP or HisTrap HP columns (GE Healthcare), depending on the purification tag. The Strep-tag binding buffer was 100 mM Tris-HCl (pH 8), 150 mM NaCl, 1 mM EDTA, 5% glycerol, 0.5 mM TCEP and elution was performed with 2.5 mM Desthiobiotin in binding buffer. The His-tag binding buffer was 25 mM Tris-HCl pH 7.4, 500 mM NaCl, 10 mM Imidazole, 10% Glycerol, 2 mM DTT and elution was performed with a four-step imidazole gradient up to 0.5M. Fractions were pooled and concentrated using 10K Amicon ultra (Milipore) followed by gel filtration on a Superdex75 16/600 column (GE Healthcare) into 20 mM HEPES pH 7.5, 300 mM NaCl, 10% Glycerol, 50 µM EDTA. After spin-concentration, aliquots were snap-frozen in liquid nitrogen and stored at -80 °C. Protein purity, quality and accurate molar mass were monitored by SDS-PAGE as well as SEC-MALS (size-exclusion chromatography multi-angle light scattering) on a Dawn Heleos II & OptiLab T-rEx (Wyatt, U.S.A.). Protein concentrations were determined on a Nanodrop spectrophotometer by absorbance at 280 nm using sequence-specific extinction coefficients^56^.

SARS-CoV-1/2 and MERS target proteins, expressed and purified from HEK293 or insect cell culture, were received lyophilized from Sino Biological (China) and dissolved to a standard concentration of 0.25 mg/mL as per manufacturer instructions, then aliquoted, snap-frozen in liquid nitrogen and stored at -80 °C. N-terminally Maleimide-labelled SpyTag peptide was synthesized by GenScript Biotech (Singapore), received lyophilized, dissolved in PBS and stored at -20 °C.

### Protein Dilutions

Target and non-target proteins were thawed on ice and centrifuged at 15.000 rpm at 4 °C for 30 to 45 min in order to remove any potential aggregates (although no aggregation was observed). Sino Biologicals proteins were then used as-is for the preparation of a dilution series starting at 320 or 640 nM. Equivalent dilutions of the Sino Biologicals storage buffer by itself did not give any sensor response. Weak background sensor signals were recorded from dilutions of DTT. In-house proteins were therefore stored or exchanged into DTT-free buffer before use. The higher-concentrated proteins from in-house production were first diluted to intermediate concentrations that could still be validated and corrected spectrophotometrically. Protein dilutions were prepared in standard sensor binding buffer (100 mM HEPES pH 7.4, 150 mM NaCl, 0.05% v/v Tween-20, 0.02% w/v NaN_3_, 0.1% w/v BSA) which was modified for saliva measurements to include cOmplete protease inhibitor cocktail with EDTA (Sigma) at 4 times the manufacturer-recommended concentration (giving a two-fold concentration in the final 1:1 mixture with saliva). BSA was not included in this saliva buffer. For measurements in the regular binding buffer, 4-fold dilution series were prepared in 96-well microplates over 23 steps starting from 320 nM. For measurements in serum, saliva and UTM, target protein dilution series were prepared in the appropriate buffer (standard or saliva binding buffer) starting from 640 nM so that final concentrations were identical after 1:1 mixture with serum, saliva or UTM.

### Surface plasmon resonance (SPR) Measurements

Surface plasmon resonance measurements were performed on a Biacore T200 instrument using Nickel-NTA sensor chips (GE Healthcare) and a modified running buffer (20 mM HEPES pH 7.4, 150 mM NaCl, 0.05% Tween-20, 0.02% w/v NaN_3_, 50 µM EDTA, prepared at room temperature and filtered) mirroring the sensor binding buffer. All analyte proteins were desalted into the running buffer (HiPrep 26/10 column, GE Healthcare) before the preparation of dilution series in the same running buffer. The GFP nanobody : GFP interaction was measured with the His_10_ -tagged nanobody immobilized (ligand) and msfGFP in solution. For the remaining measurements, since all viral target proteins arrived with a non-cleavable His-tag, target proteins were immobilized instead of the nanobody. All ligand proteins gave stable immobilization responses with minimal signal loss over time. His tags were removed from nanobody-spyCatcher proteins by overnight cleavage with excess 3C protease (produced in-house), followed by gel filtration on a Superdex200 Increase 10/300GL column. All ligand proteins were immobilized to equal loading levels of around 100 RU at 10 µl/min flow rates. Binding and unbinding experiments were run at a flow rate of 80 µl/min. Two replicates were prepared of each dilution series and measured in the course of the same experiment. Between each measurement cycle, the sensor surface was regenerated with 0.35 M EDTA in running buffer and re-charged with 0.5 mM NiSO_4_ in water. Biacore results were analyzed with the manufacturer analysis software version 2.1 following standard procedures (double-subtraction of reference channel and buffer injection signal) applying a 1:1 binding model and simultaneous curve-fitting. SARS-CoV-2 RBD results were instead fit a heterologous ligand binding model.

### Fluorescence imaging

The GFP nanobody functionalized gold electrodes before and after its incubation with a GFP solution together with the negative controls (the biofunctional electrode before and after incubation with mCherry) were imaged using fluorescence microscopy. Electrodes were placed between two cover slips in the presence of PBS in order to keep the flexible material in plane and in focus. Pictures were recorded on a fluorescent inverted microscope DMI8 (Leica Microsystem) coupled with pE-4000 fluorescence illumination system (CoolLED).

### OECT characterization and sensor operation

We used a Keithley source meter to apply drain (*V*_D_) and gate voltages (*V*_G_) and obtain gate and channel currents (*I*_G_ and *I*_D_) in ambient conditions. A PDMS well was glued on top of the channels and filled with 75 µL of 10 mM PBS. The steady-state transistor characteristics were obtained by measuring *I*_D_ vs. *V*_D_ at various *V*_G_, for PEDOT:PSS applied between -0.6 V to 0.6 V with 0.1 V step (2.5 V/s) and for p(g0T2-g6T2) from 0.2 V to -0.6 V with 0.05 V step (1.25 V/s). *V*_*D*_ was swept from 0 V to -0.6 V. We first chose a channel and obtained its transfer curve (*I*_D_ vs. *V*_G_) in PBS using the nanobody-functionalized gate electrode incubated for 10 min in the buffer solution (buffer, saliva, UTM or serum, in the absence of target proteins). The currents obtained were used as the baseline signal (*I*_0_). The same gate electrode was then incubated with 5 µL of the solution (buffer, saliva, UTM or serum) that contains the protein target for 10 min (pipetting 30 s every 3 min). The electrode was washed thoroughly with PBS to remove any unbound proteins. To obtain calibration curves, we prepared various concentrations of the target protein and collected device data starting from the most diluted one. The normalized response (NR) of the sensor was calculated according to the following equation:

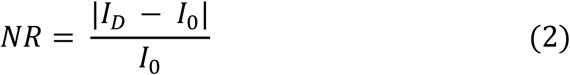

where *I*_*D*_ is the current response of the sensor to an analyte solution that the gate was exposed to. According to IUPAC, the LOD was calculated as the concentration leading to a response that equals to the average of the noise level plus three times the noise standard deviation^57,58^. The level-of-noise 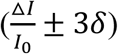 was taken from the relative current variation in negative control sensors.

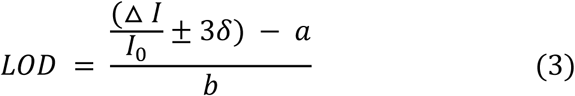

where 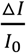 is the average response of the blank sample, *δ* is the relative standard deviation, *a* and *b* are the intercept and slope of the calibration curve, respectively.

### Clinical Sample Preparation and Testing

The samples (saliva samples and nasopharyngeal swabs) used in this study (Fig. 5) were collected from human subjects as part of registered protocols approved by the Institutional Review Board of King Faisal Specialist Hospital & Research Center and KAUST Institutional Biosafety and Bioethics Committee (IBEC). All volunteers provided signed consent to participate in the study. Nasopharyngeal swabs collected from COVID-19 patients and healthy subjects were stored in UTM, and saliva was collected inside empty urine beakers or 50 mL tubes. Samples were stored at 4 °C and tested within one to two days after collection using the same protocol described above. The saliva used for original sensor characterization with recombinant proteins was self-collected in the morning before food or tooth brushing, filtered and stored in aliquots at -20 °C. All volunteers provided signed consent to participate in the study and provide saliva. Human serum from human male AB plasma was purchased from Sigma-Aldrich and used as-received. All protocols and procedures involving human saliva, nasopharyngeal swabs and serum were approved by the KAUST IBEC under approval number 18IBEC11 and 20IBEC25, NCBE, KSA registration number HAP-02-J-042.

### Data and Materials availability

All the data needed to evaluate the conclusions in the paper are present in the paper and/or the Supplementary Materials. Additional data related to this paper may be requested from the authors.

## Supporting information

Guo et al. Supplemental Information

## Acknowledgements

Fig. 1 was produced by Xavier Pita, scientific illustrator at KAUST. The authors thank all members of the KAUST Rapid Research Response Team (R3T) for COVID-19, especially Prof. Samir Hamdan for their contributions in this study. The authors convey thanks to Sara Mfarrej, Amit K. Subudhi, and Prof. Arnab Pain for providing access and assisting the experiments in the BSL 2 plus category experimental room at KAUST. We thank the KAUST Health team (operated by Soliman Fakeeh Hospital, Jeddah), including Daniel Buttigieg for providing clinical samples. We thank staff in King Faisal Specialist Hospital and Research Center (Riyadh), particularly Dr. Ali Alzahrani, Madain Alsanea and Faten Alhadeq, for their help in organizing and hosting the clinical studies. The authors thank KAUST nanofabrication core lab team, Diego Rosas Villalva and Dr. Ullrich Buttner for their help in device fabrication and integration. This work was initiated thanks to the KAUST Impact Acceleration Fund (IAF) program. The research reported in this publication was supported by funding from King Abdullah University of Science and Technology (KAUST), Office of Sponsored Research (OSR), under award numbers REI/1/4204-01, REI/1/4229-01, OSR-2018-CRG7-3709, OSR-2018-CARF/CCF-3079, OSR-2015-CRG4-2572 and OSR-4106 CPF2019. We acknowledge EC FP7 Project SC2 (610115), EC H2020 (643791), and EPSRC Projects EP/G037515/1, EP/M005143/1, and EP/L016702/1.

## Author information

These authors contributed equally: Keying Guo, Shofarul Wustoni and Anil Koklu.

## Contributions

S.I., S.A., and R.G. conceived the research, designed the experiments, supervised the work and wrote the manuscript. K.G, S.W., and A.K. fabricated the devices and performed the OECT experiments. K.G and S.W. functionalized the gate electrodes. A.K. performed EIS measurements, S.W. performed CV and XPS measurements, and K.G. conducted QCM-D experiments. R.G. and E.D.G. designed and produced recombinant proteins and performed Biocore experiments. A.H. took the fluorescence microscope images and developed the LabView codes to operate the OECTs. A. N. A. collected clinical samples and provided RT-PCR results. F. S. A. supervised the clinical experiments with A. A. A. and R.G. and conducted RT-PCR. M. M. and I. M. provided the p-type material. All the authors were involved in the discussion and participated in manuscript input.

## Competing Interests

A US provisional application (no. 63/047,547 -“Nanobody Functionalized Electrochemical Transistors and Methods of Making and Using Thereof”) related to this work was filed by S.I, S.A., R.G., K.G., S.W., and A.K.

## Notes

### Author Declarations

protocols were approved by the Institutional Review Board of King Faisal Specialist Hospital & Research Center and KAUST Institutional Biosafety and Bioethics Committee (IBEC). All protocols and procedures involving human saliva, nasopharyngeal swabs and serum were approved by the KAUST IBEC under approval number 18IBEC11 and 20IBEC25, NCBE, KSA registration number HAP-02-J-042.

