## Supplementary material for "A nanobody-functionalized organic electrochemical transistor for the rapid detection of SARS-CoV-2 and MERS antigens at the physical limit": Guo et al. Supplemental Information

### 1. Further discussion on XPS data

The S 2p XPS spectra presented in **Figure 2b** show three deconvoluted peaks with a binding energy at 161.4, 162.7, and 164.1 eV for the HDT-functionalized gold electrode. The peaks at 161.4 and 162.7 eV stem from the sulfur chemisorbed on the gold surface through a thiolate bond (Au-S)[1,2] . The resolution of these peaks indicate the high-quality HDT formed on the surface. The signal at 164.1 eV, on the other hand, corresponds to the free thiol group (terminal R-SH), suggesting two types of sulfur species present in the monolayer; thiolate-type sulfur (Au-SR) and tail thiol sulfur (R-SH). These results suggest that the HDT molecules are in a standing-up configuration with an upright molecular structure. They are bound to gold via the thiolate link using one of their thiol groups, while the other thiol group is free and located at the SAM-air interface. If the HDT molecules were present exclusively in a lying-down configuration, we would be only detecting thiolate components, as all S atoms would be thiolate linked to the gold surface. As we introduced the SpyTag peptide and nanobody-SpyCatcher on this surface, a new peak appeared at ca. 163 eV, originating from the Sulphur in methionine[3].

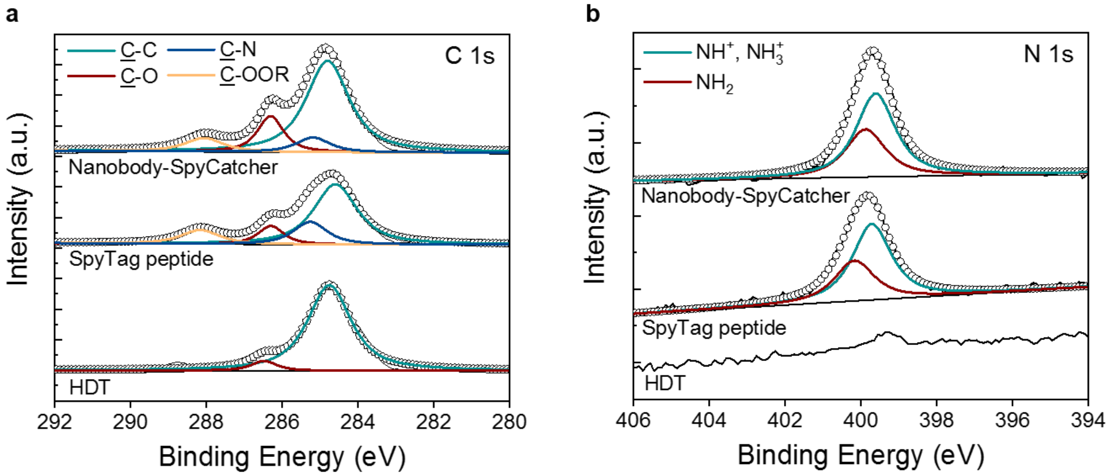


**Figure S1. Surface characterization of the Au electrode upon Chem-SAM and Bio-SAM immobilization.** High-resolution of **a)** C 1s and **b)** N 1s XPS spectra of the gold electrode after HDT, SpyTag peptide, and the nanobody-SpyCatcher protein functionalization. The nanobody-SpyCatcher buffer contained BSA.

In the high-resolution of C 1s **(Figure S1a)**, we detect the C-C peak of the HDT-functionalized gold, attributed to the six-carbon chain of the HDT molecule. The C-O, C-N and C-OOR peaks originate from the amino acids in SpyTag peptide and the nanobody-SpyCatcher. The N 1s spectra **(Figure S1b)** show that nitrogen groups appear on the gold surface upon the introduction of the Bio-SAM proteins.

### 2. Cyclic voltammetry curves and electrochemical impedance spectroscopy

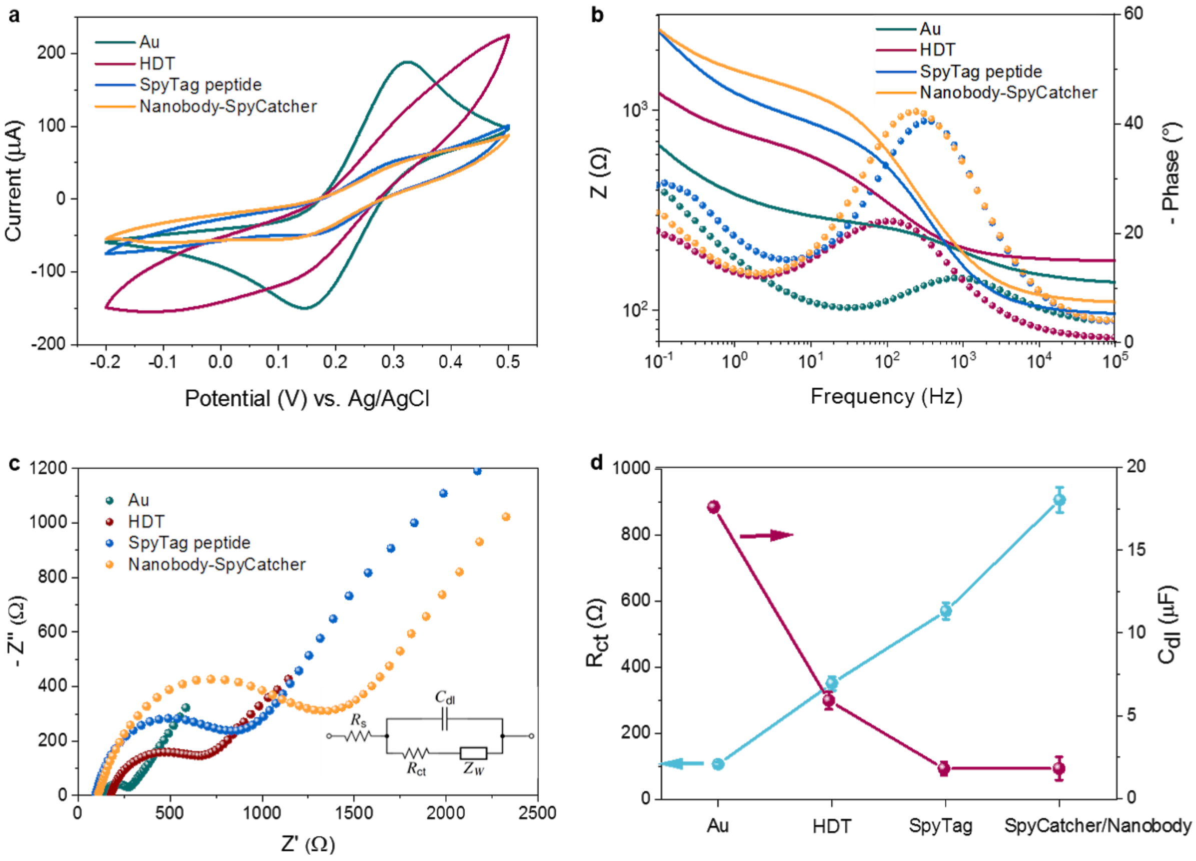


**Figure S2. Electrochemical characterization of the biofunctionalized Au electrode. a)** Cyclic voltammogram, **b)** Bode plot (solid lines and dotted lines are corresponding to the magnitude and the phase of the impedance, respectively), and **c)** Nyquist plot of the gold electrode before and after the subsequent functionalization with HDT, SpyTag peptide, and the nanobody-SpyCatcher. Inset in **c)** is the equivalent circuit model used to fit the impedance spectra. **d)** The calculated $R_{ct}$ and $C_{dl}$ changes of the Au electrode.

The gold electrode exhibits the well-known reversible peaks for the $[Fe(CN)_6]^{3-/4-}$ redox couple with a peak potential separation of approximately 180 mV (**Figure S2a**). After the modification with HDT, the electron transfer of the redox couple is inhibited, suggesting that a continuous HDT layer has blocked the gold electrode surface. When the SpyTag peptide and nanobody-SpyCatcher are immobilized on the HDT assembled gold electrode, the electrical communication of the Au layer with the redox probe is further suppressed as indicated from the reduced current values. **Figure S2b and c** show the representative Bode and Nyquist plots of the Au gate electrode after each functionalization step. We used the Randles equivalent circuit model to quantitatively analyze the impedance spectra (inset of Figure S2c). It consists of the electrolyte resistance ($R_s$), electric double layer capacitance ($C_{dl}$) formed at the electrode/electrolyte interface, charge transfer resistance ($R_{ct}$) of the electrode, and Warburg impedance resulting from the diffusion of ions from the electrolyte to the electrode surface. As HDT was assembled, the diameter of the semicircle in the high frequency region of the Nyquist trace increased significantly, indicating an increase of impedance. Modeling the Nyquist plots suggests that the $R_{ct}$ increases from 0.1 to 0.4 kΩ with HDT addition, confirming the charge blocking behavior of the HDT (**Figure S2d**). The impedance further increases and the Nyquist plot becomes a large semicircle that extends across the entire range of frequencies after the immobilization of SpyTag peptide ($R_{ct} \approx 0.6$ kΩ) and SpyCatcher/nanobody($R_{ct} \approx 1$ kΩ). In **Figure S2d**, we also summarize the effect of SAMs on $C_{dl}$, which shows a monotonic decrease after each functionalization step, consistent with the trend observed in CV curves. These measurements suggest that the SAMs reduce the electrochemical capacitance and increase the charge transfer resistance of the Au electrode, verifying their successful immobilization on the surface.

### 3. Further discussion on QCM-D data

Operating as a very sensitive balance, QCM-D can quantitatively monitor the real-time changes in the mass of the Au electrode upon each biofunctionalization step, i.e., the SpyTag coupling followed by the nanobody-SpyCatcher capture.[7] When mass (e.g. peptide, nanobody, BSA) is accumulated on the gold sensor, we observe a decrease in its oscillation frequency. As shown in **Figure 2c and S3**, the frequency (top panel) decreases and the mass density (bottom panel) increases upon injection of SpyTag peptide and nanobody-SpyCatcher on the HDT coated surface. The mass density increased to 111 ng cm$^{-2}$ after SpyTag-peptide attachment through maleimide conjugation between sulfhydryl groups of the HDT and maleimide groups of the peptide. The SpyTag sequence on the peptide can specifically couple with the SpyCatcher domain linked to the nanobody. We performed two measurements with two binding buffers. One buffer contained only the nanobody-SpyCatcher (**Figure 2c**). In a second experiment we used the final formulation of sensor binding buffer comprising also the BSA (**Figure S3**). After thorough rinsing with PBS, a mass density of 406 ng $cm^{-2}$ was gained with nanobody-SpyCatcher, corresponding to $8.6 \times 10^{12}$ nanobodies per $cm^2$. A larger mass density of 622 ng $cm^{-2}$ was obtained when the immobilization solution also contained BSA. These results suggest two scenarios or a combination of them: 1) BSA binds to and blocks: sulfhydryl groups exposed to the solution, gaps between peptide linkers or other surface imperfections. 2) BSA acts as a blocking agent *in solution* by capturing contaminating or partially unfolded proteins so that only intact nanobody-SpyCatcher fusions can couple to the surface. In support of scenario (2), switching to PBS does not cause any visible detachment of  unbound molecules when BSA was added to the immobilization solution (unlike in the non-BSA immobilization trace shown in the main manuscript Figure 2d). Moreover, adding BSA directly to the immobilization solution outperformed protocols using a separate BSA blocking step.

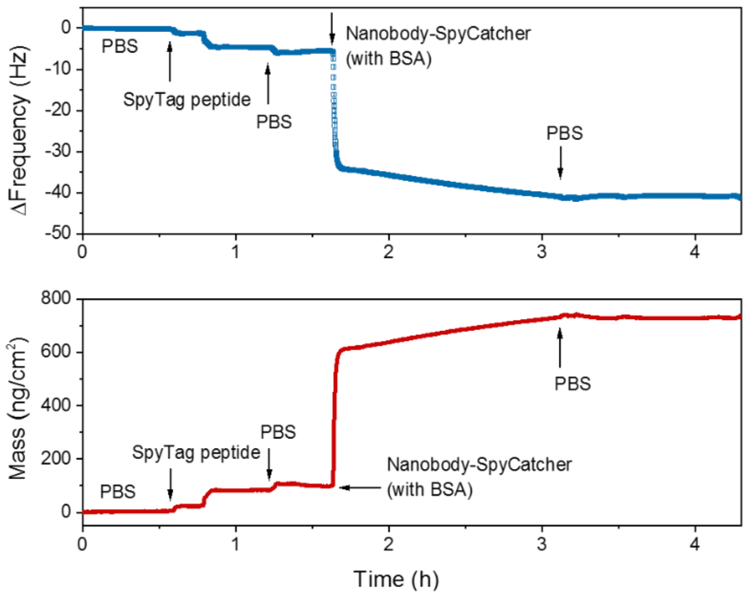


**Figure S3. Monitoring of the biofunctionalization of the gold surface using QCM-D.** The gold QCM-D sensor comprising an HDT SAM was subjected to the two-step functionalization protocol consisting of the SpyTag peptide coupling followed by the nanobody-SpyCatcher immobilization. In this case, nanobody-SpyCatcher solution also contained BSA as a blocking agent. The top panel shows the change in QCM-D frequency of the sensor ($7^{th}$ overtone) over time as SpyTag peptide, PBS, nanobody-SpyCatcher and PBS was introduced to the system. The bottom panel presents the corresponding mass changes that the sensor undergoes during these steps. PBS was used to wash away the unbound molecules.

### 4. Optimization of the OECT biosensor geometry

The sensor response was optimized by tuning the geometry of the gate electrode and the channel. Three differently sized gate electrodes (0.8 mm in square, 2.8 mm in circle, and 4.8 mm in square) were fabricated to gate OECT channels with two distinct geometries where we varied the channel length, $L$ = 10 µm or $L$ = 100 µm, while keeping the channel width constant at 100 µm. The gate electrodes were functionalized with the GFP nanobody and sensor performance was evaluated as shown below. The smaller gate and channel exhibited the highest change in current upon GFP binding.

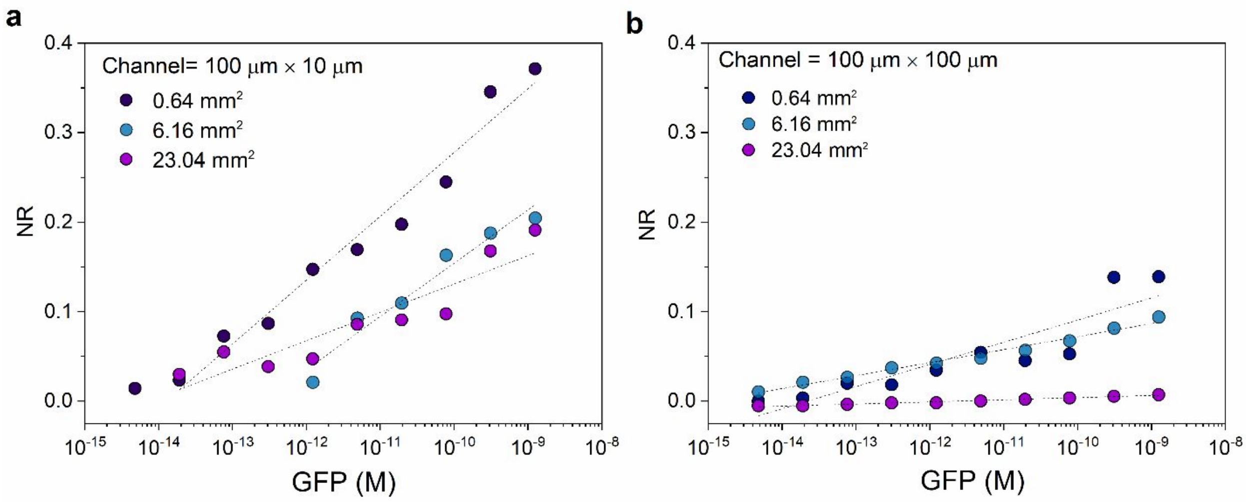


**Figure S4. The effect of channel and gate geometry on the performance of PEDOT:PSS OECT sensors.** The GFP-functionalized Au electrodes with an area of 0.64 mm$^2$ (0.8 × 0.8 mm), 6.16 mm$^2$ (diameter = 2.8 mm), and 23.04 mm$^2$ (4.8 × 4.8 mm) are used to gate **a**) channel with a width ($W$) 100 µm and length ($L$) 10 µm, and **b**) channel with a $W$= 100 µm and $L$= 100 µm.

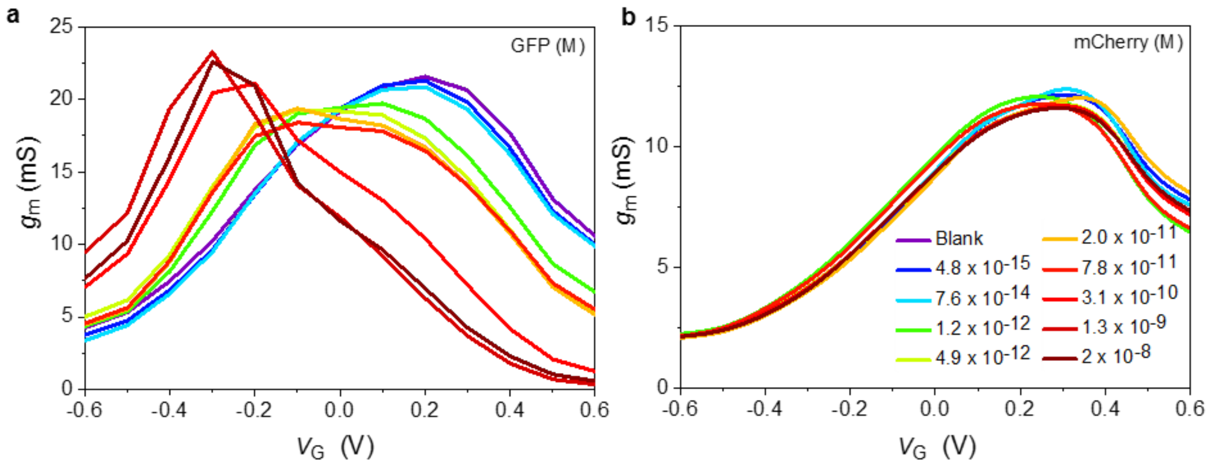


**Figure S5. Transconductance *vs* gate voltage for the GFP nanobody-functionalized Au electrode gating a PEDOT:PSS channel.** The protein in the incubation buffer was **a)** GFP and **b)** mCherry. The protein concentration ranges from 5 fM to 20 nM. The purple curve corresponds to the gate electrode incubated in the buffer only (blank). The $V_G$ that attains the max $g_m$ shifts from ca. 0.3 V to -0.3 V with GFP binding.

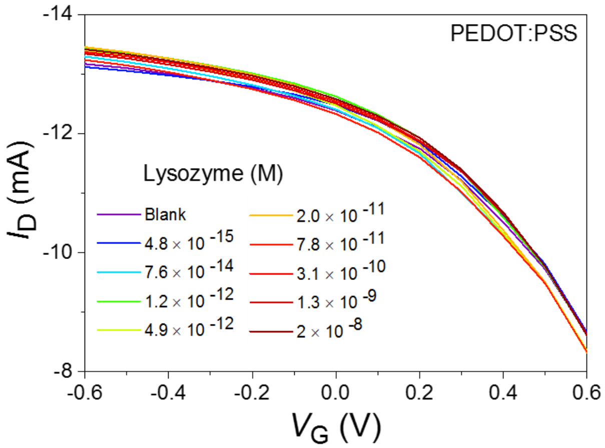


**Figure S6. Sensor response to non-target proteins.** Transfer characteristics of the GFP nanobody-functionalized Au electrode gating a PEDOT:PSS channel as the gate is exposed to various concentrations of lysozyme.

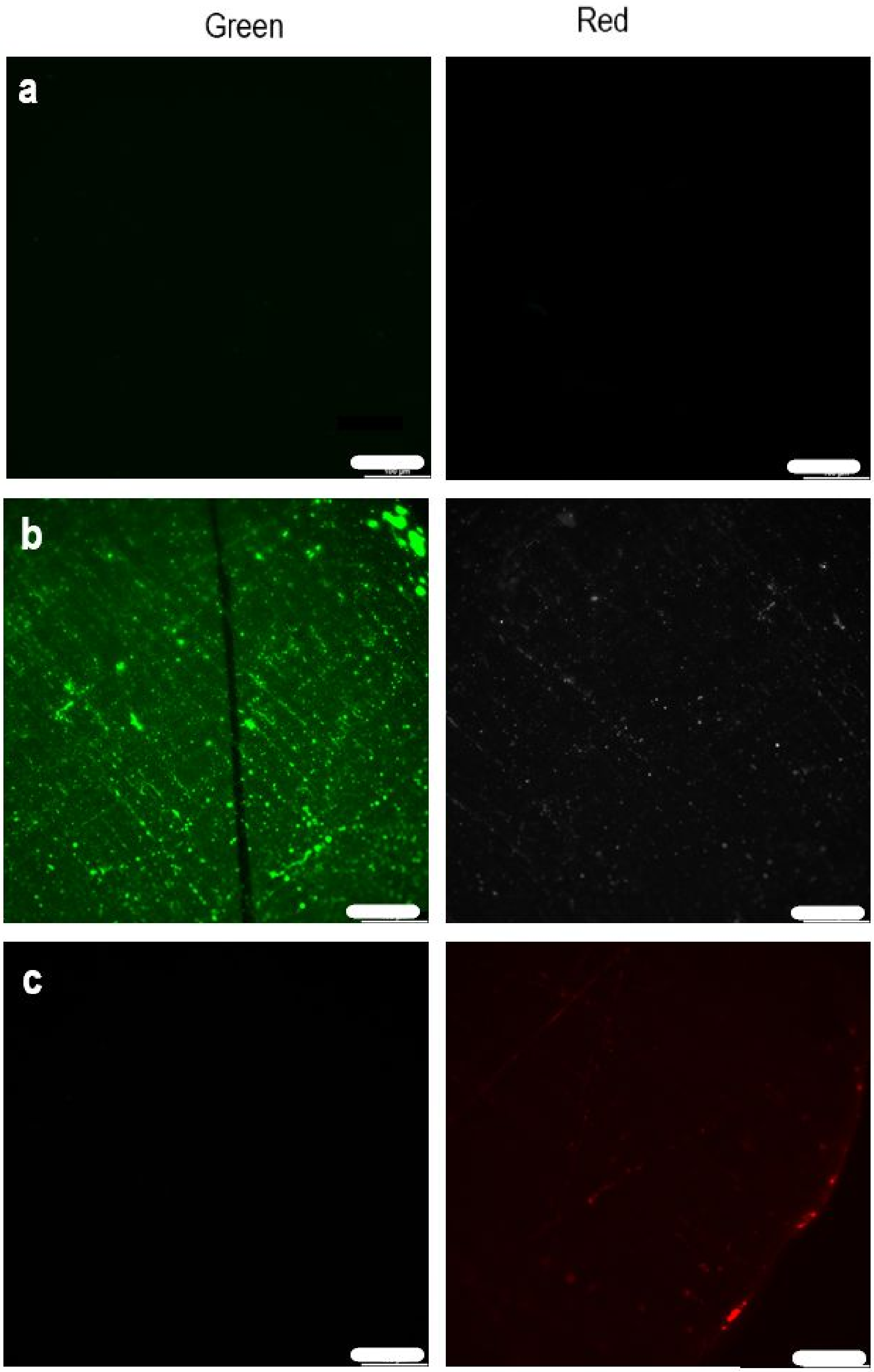


**Figure S7. Fluorescence images of the GFP nanobody-functionalized Au electrode.** The biofunctionalized gate electrodes are shown **a)** in PBS (blank), **b)** after incubation with GFP (20 nM), **c)** after incubation with mCherry (20 nM). The green is the fluorescence from the surface bound GFP and the red at the edge of the gate electrode is the fluorescence from adsorbed mCherry. Scale bar is 100 µm. The electrode surface was scratched on purpose using a tweezer, which is the dark line seen in b), in order to gain color contrast.

As GFP is bound to the nanobody functionalized Au electrode, the film, which is originally nonfluorescent at the chosen wavelength (Figure S7a, left) emits green fluorescence (Figure S7b, left). As the electrode is incubated in the mCherry solution, only a few spots on the edge of the sensor show red fluorescence (Figure S7c, right). This signal is caused by the mCherry which can be non-specifically absorbed, mostly on the rough or damaged edges of the surface resulting from the laser cutting process.

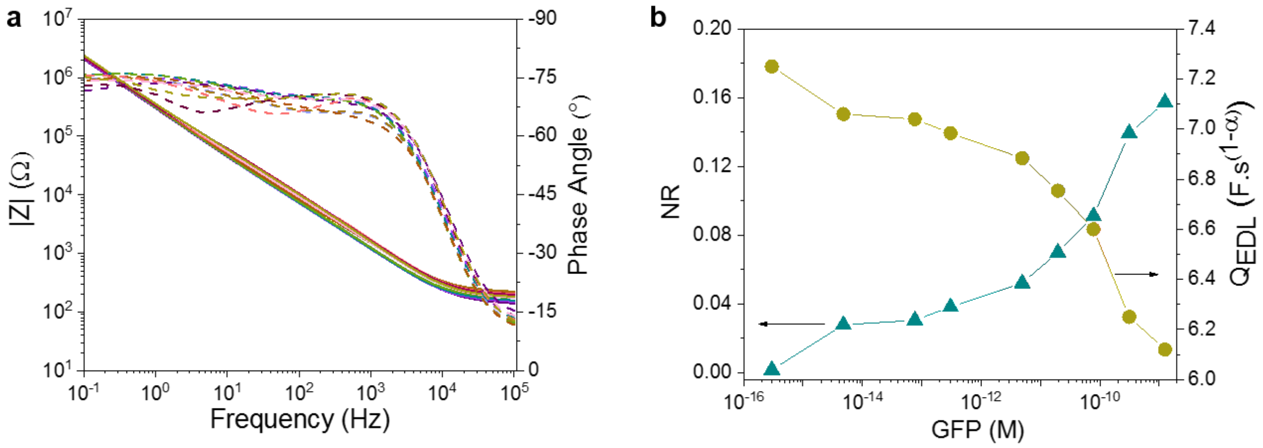


**Figure S8. Electrochemical impedance spectra of the GFP nanobody-functionalized electrode upon GFP binding. a)** Bode plots of the electrode upon successive incubations in increasing GFP concentrations **b)** the normalized change in in the corresponding constant phase element component obtained by equivalent circuit model analysis.

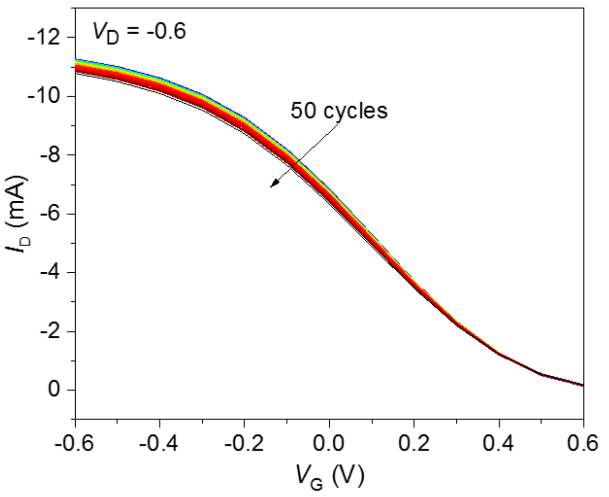

**Figure S9. The cycling stability of a Au electrode gated OECT comprising a PEDOT:PSS channel**. The decrease in the drain current is ~3% at $V_D = V_G$ = -0.6 V upon 50 I-V cycles.

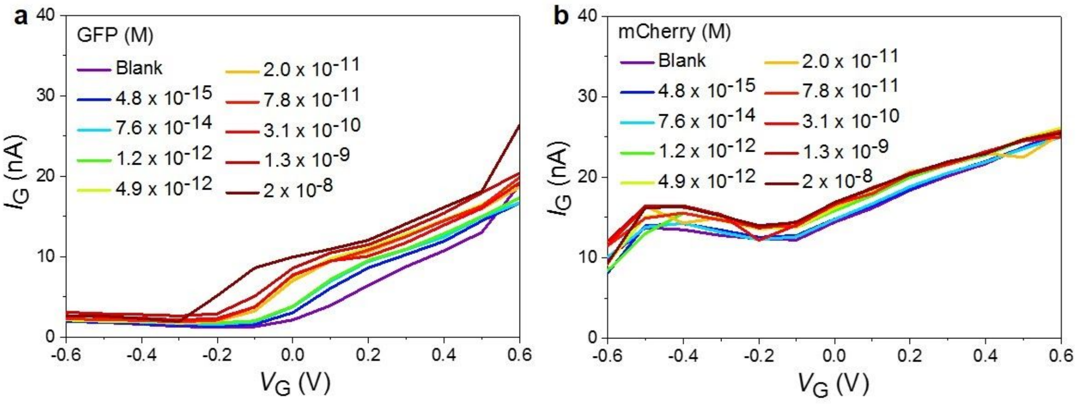


**Figure S10. The gate currents of the GFP biosensors**. The changes in the gate current ($I_G$) of the GFP nanobody-functionalized OECT as the gate is exposed to various concentrations of **a)** GFP and **b)** mCherry. The channel is PEDOT:PSS.

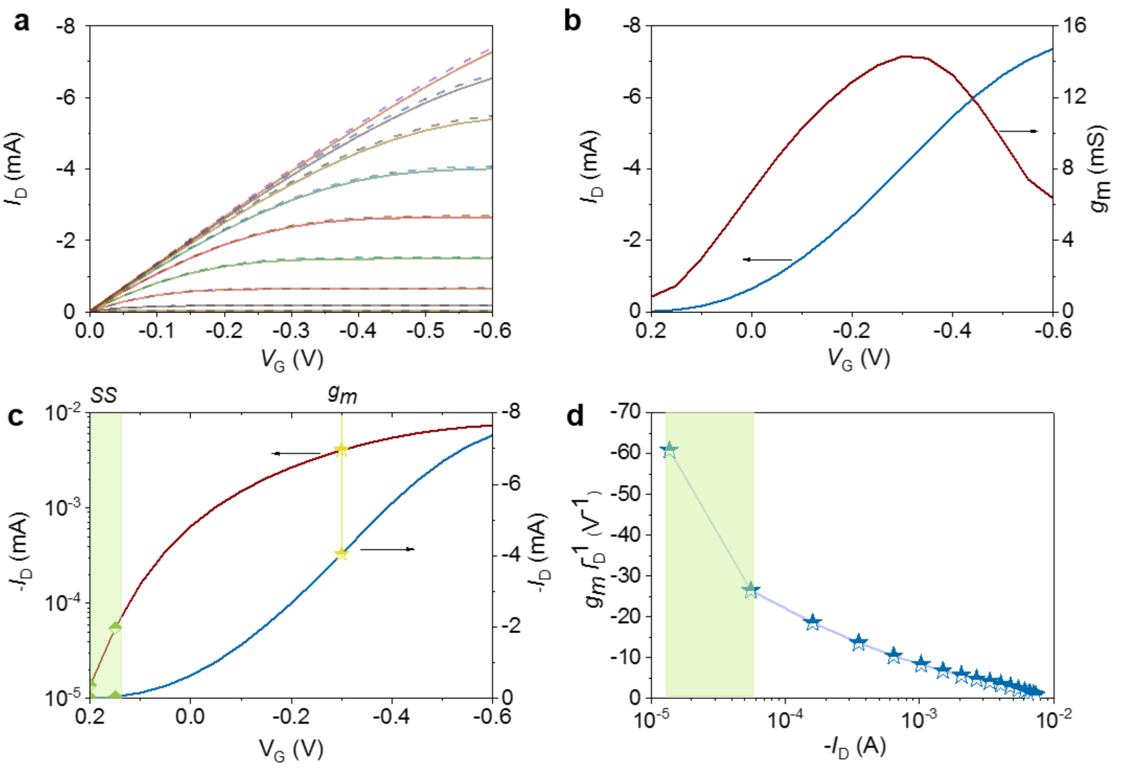


**Figure S11. Steady-state characteristics of p(g0T2-g6T2) OECTs**. **a)** Output curves ($I_D$ *vs.* $V_D$) and **b)** the transfer curve (left axis) with the corresponding transconductance ($g_m$ *vs.* $V_G$, right axis). $V_D$ was -0.6 V. **c)** Transfer characteristics in the linear (right axis) and log scale (left axis). The operating points of peak transconductance ($g_m$, yellow stars) and peak subthreshold slope (SS, region highlighted in green) are marked. **d)** Transconductance efficiency ($g_m$/ $I_D$) *vs.* $I_D$ for $V_D$=-0.6 V.

Figure S11 a-b shows typical output and transfer curves of a Au electrode gated p(g0T2-g6T2) channel. The slope of the transfer curve represents the transconductance, $g_m$, and exhibits a peak value of 15 mS at $V_G$ = -0.3 V. The subthreshold region can be seen in the log scale plot of $I_D$ *vs.* $V_G$ with the maximum slope of 60 mV dec$^{-1}$, at $V_G \approx 0.15$ V (**Figure 11c**). The devices have the highest gain and efficiency at low $V_G$ range (**Figure 11d**).

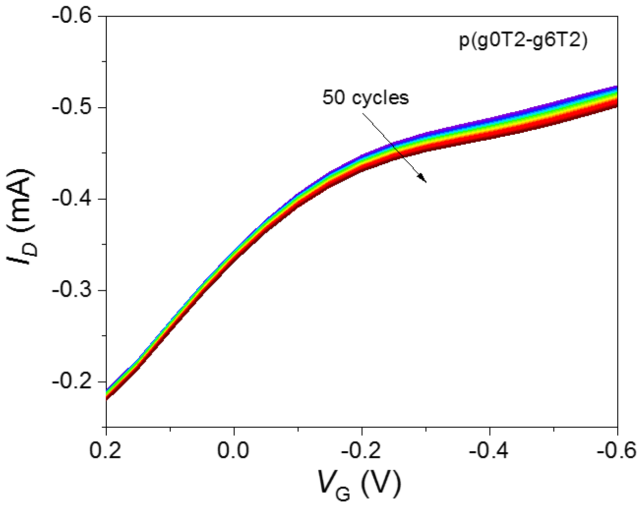


**Figure S12. The cycling stability of a Au electrode gated OECT comprising a p(g0T2-g6T2) channel**. The decrease in the drain current is ~1% at $V_D = V_G = -0.1$ V upon 50 I-V cycles.

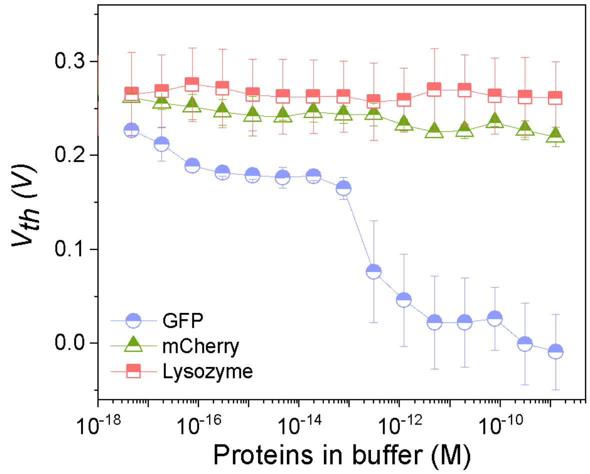


**Figure S13. The change in the threshold voltage ($V_{th}$) upon protein detection.** The changes in $V_{th}$ of p(g0T2-g6T2) OECTs as the gate electrode is exposed to GFP, mCherry and Lysozyme at various concentrations. The $V_{th}$ shifts towards more negative values as more GFP molecules bind to the gate electrode. Error bars represent the standard deviation from at least three gate electrodes.

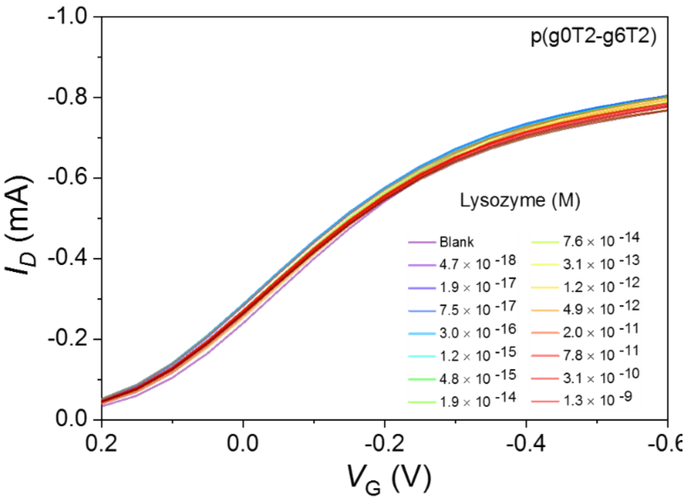


**Figure S14. Sensor response to non target proteins.** Transfer characteristics of the GFP nanobody-functionalized Au electrode gated p(g0T2-g6T2) channel as the gate is exposed to various concentrations of lysozyme.

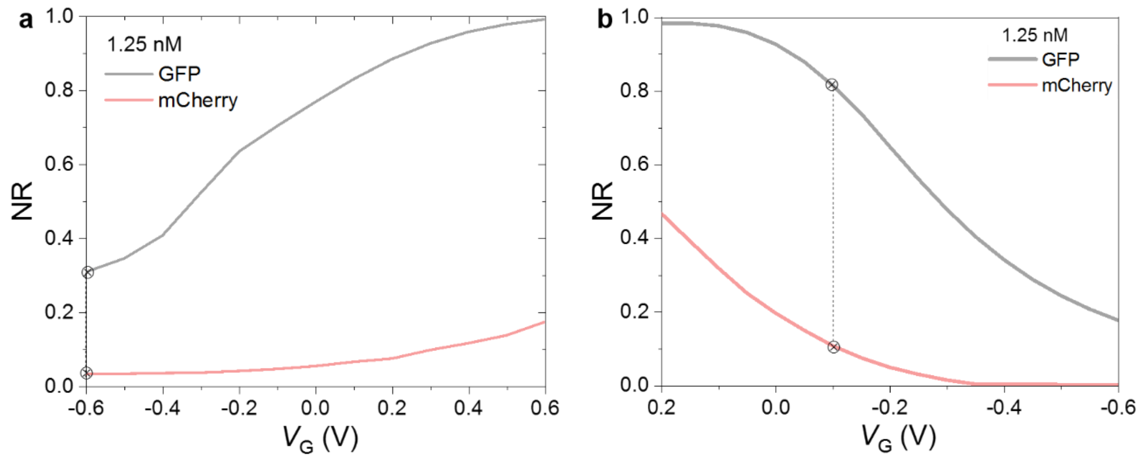


**Figure S15. The gate voltage dependence of sensor response.** The normalized response curves as a function of $V_G$ for OECTs comprising **a)** PEDOT:PSS **b)** p(g0T2-g6T2). The gate electrode was incubated in 1.25 nM of protein solution. $V_D$ is -0.1 V and -0.6 V for p(g0T2-g6T2) and PEDOT:PSS, respectively. Although the NR is maximized at $V_G$ = 0.6V for PEDOT:PSS, since the polymer is almost fully de-doped at this biasing condition (depletion mode), the sensor current response could not be well-resolved. We therefore chose to calculate NR at $V_G$=-0.6V.

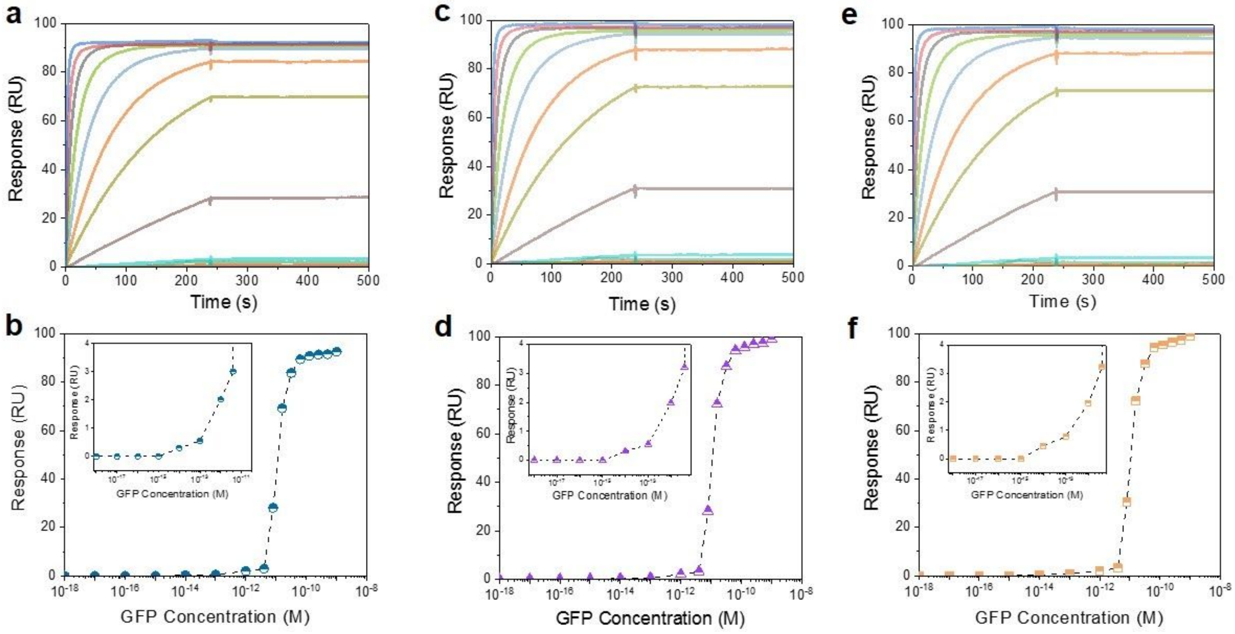


**Figure S16. SPR measurement of the GFP : nanobody interactions.** His-tagged GFP nanobody was immobilized with a low loading level (**a**) and, for comparison of SPR and OECT sensitivities, with a high loading level (**c**, **e**) on a Ni-NTA sensor chip and exposed to increasing concentrations of GFP. The kinetic off rate was too low to be quantified, indicating exceptional stability over time of the complex once formed. Analysis of steady-state binding levels (**b**, **d**, **f**) indicated first SPR signals for GFP concentrations as low as 1 pM ($10^{-12}$ M).

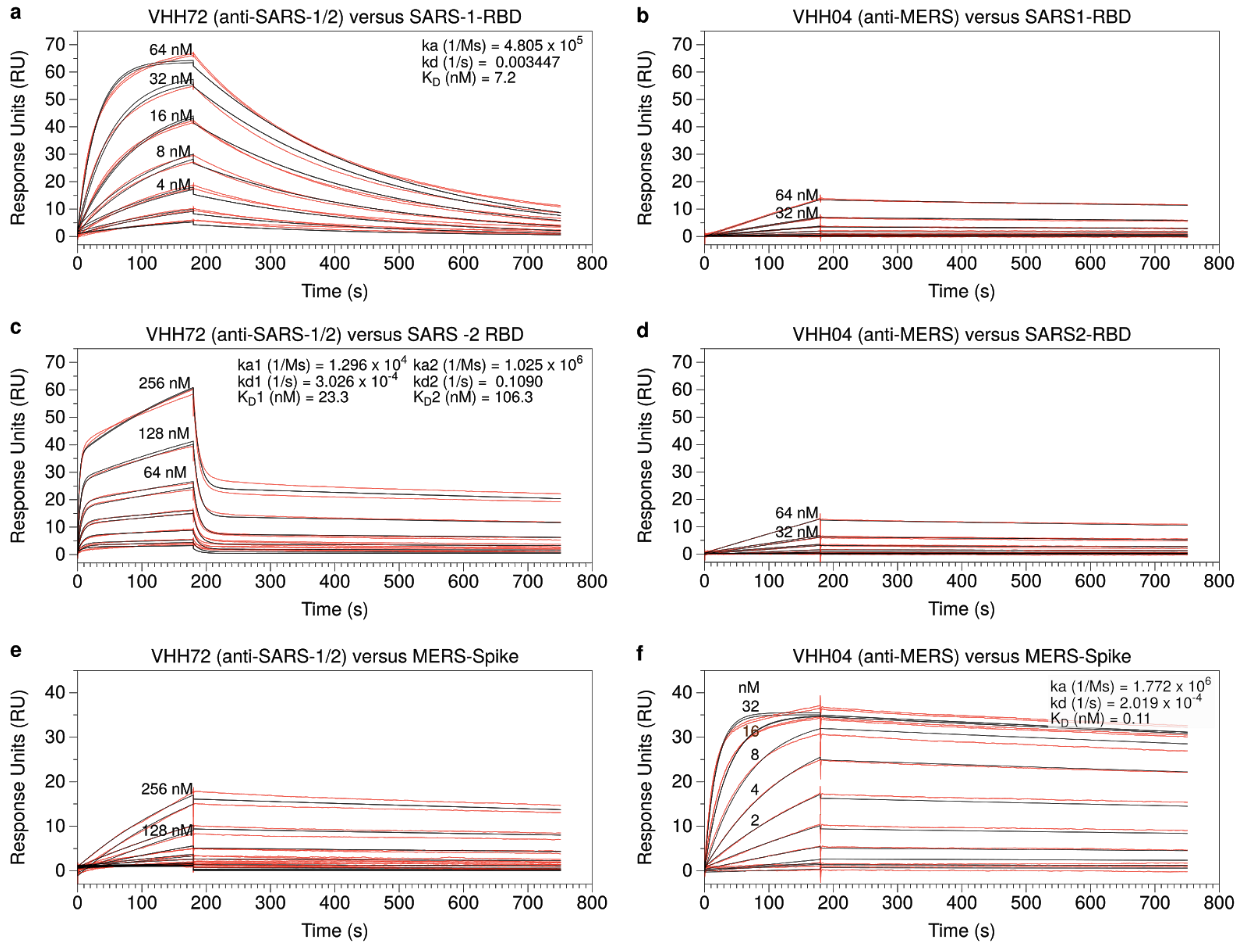


**Figure S17. SPR measurements of anti-SARS and anti-MERS nanobodies.** Three different viral proteins (SARS-CoV RBD, SARS-CoV-2 RBD, MERS-CoV S1) were immobilized to similar loading levels in three SPR channels and exposed (in parallel) to increasing concentrations of anti-SARS (VHH72) or anti-MERS (VHH04) nanobody. On- and off-rates and kinetically determined equilibrium dissociation constants ($K_D$) are given. Red traces: SPR response (after double-subtraction of reference channel and buffer injection signal) from two replicate dilution series', black traces: multi-curve kinetic model fit. The interactions of VHH72 with SARS-1 RBD as well as that of VHH04 with MERS Spike protein were fit with a 1:1 binding model. The interaction between VHH72 and SARS-CoV-2 was fit with a heterologous ligand binding model indicating a majority population of slow on, slow off- and a minority population of fast on, fast off binding events. (**b, d**) A weak cross-reaction is suggested for VHH04 binding off-target to SARS-CoV RBD but would require far higher analyte concentrations for quantitative confirmation. (**e**) potential off-target binding of VHH72 to MERS-CoV cannot be ruled out but would be weaker by comparison (Note that maximum analyte concentrations are higher than in b and d).

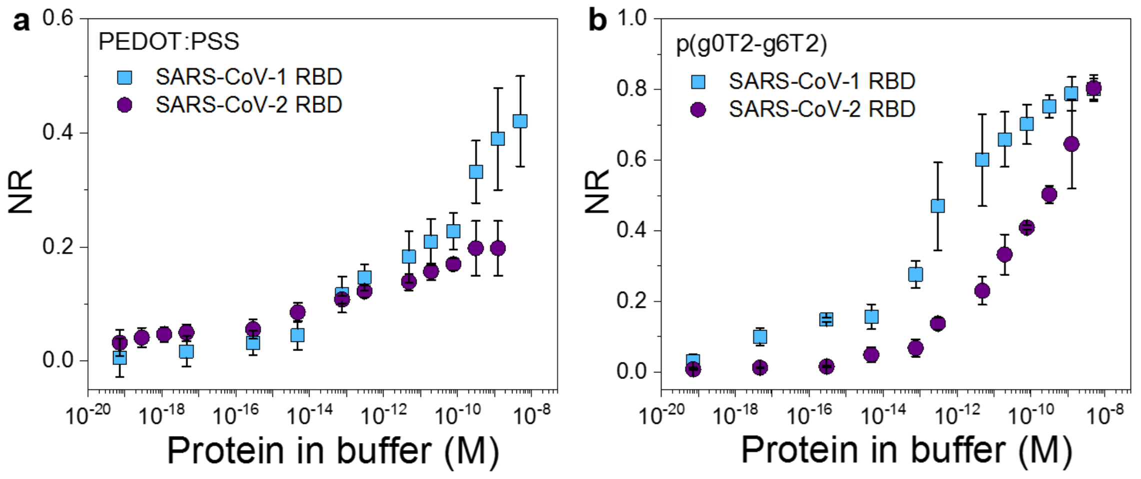


**Figure S18. The SARS-CoV nanobody functionalized OECT response to SARS-CoV RBDs.** Normalized response (NR) of the SARS-CoV nanobody-functionalized electrodes gating **a)** PEDOT:PSS and **b)** p(g0T2-g6T2) channel. The electrodes were exposed to various concentrations of SARS-CoV(1) RBD or SARS-CoV-2 RBD in the buffer. p(g0T2-g6T2) OECT has an LOD of $1.6 \times 10^{-17}$ M for SARS-CoV(1) RBD; the PEDOT:PSS OECT gives an LOD of $1.5 \times 10^{-15}$ M.

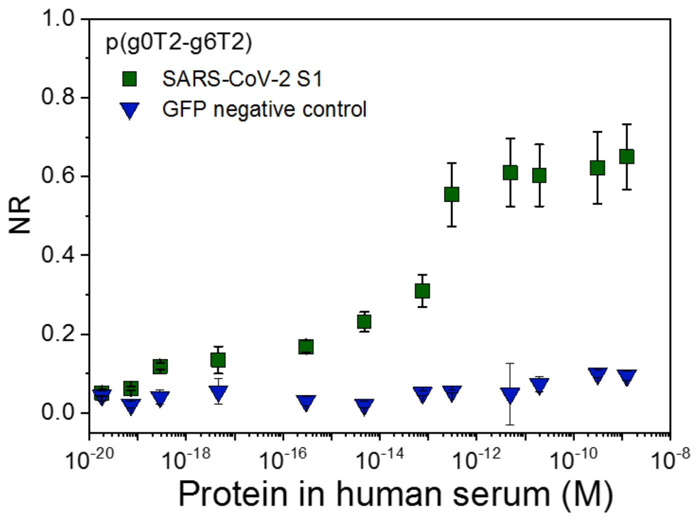


**Figure S19. The OECT biosensor response to SARS-CoV-1 RBD in human serum.** The normalized response (NR) of SARS-CoV-1 nanobody-functionalized electrode gating a p(g0T2-g6T2) channel to SARS-CoV-2 S1 and GFP spiked in human serum.

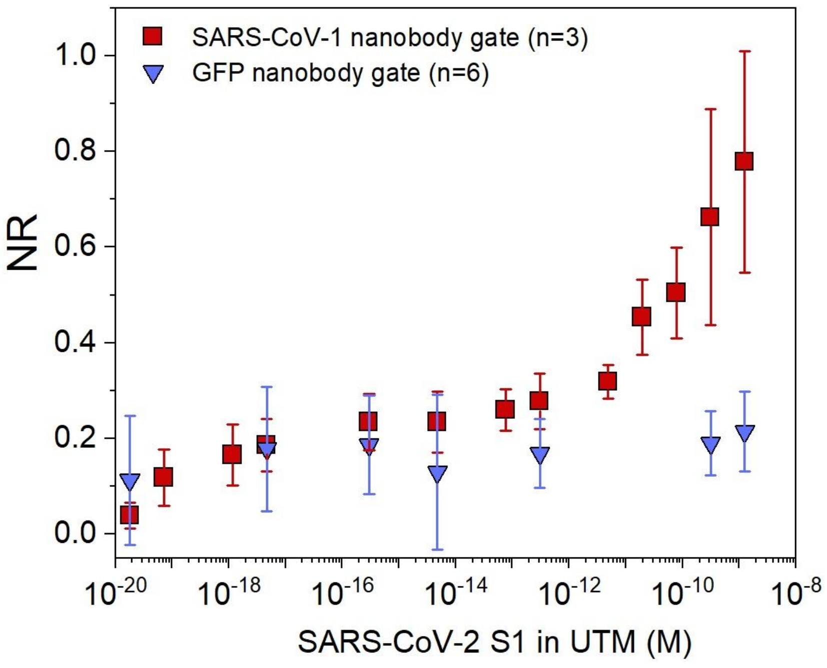


**Figure S20. The OECT biosensor response in UTM.** The response of nanobody-functionalized OECTs comprising p(g0T2-g6T2) channel to SARS-CoV-2 S1 protein spiked in UTM.

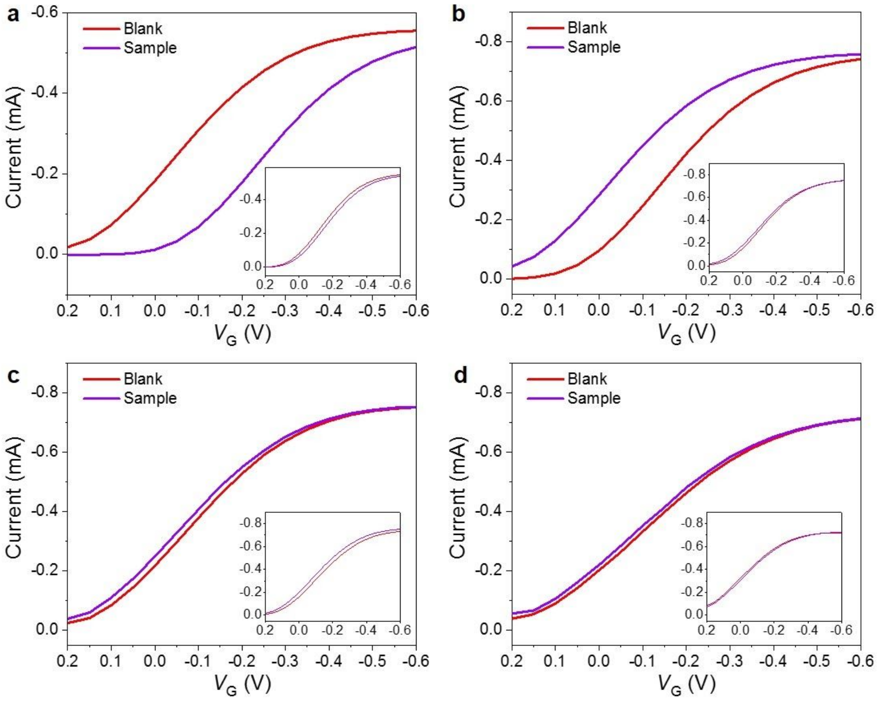


**Figure S21. Exemplary OECT biosensor response to patient samples. a)** Nasal swab sample 10, **b)** Saliva sample 13, **c)** Nasal swab sample 4, **d)** Saliva sample 4. The inset figures were obtained using the GFP nanobody gate. Nasal swab sample 10 and Saliva sample 13 were confirmed to be positive, while Nasal swab sample 4 and Saliva sample 4 were confirmed to be negative by RT-PCR.

### Table S1. LOD of the OECT biosensors.

| Channel Material | Nanobody on the Gate | Targets | Sensors in Buffer | | |
|---|---|---|---|---|---|
| | | | LOD++ | Fitting Equation | Highest SD |
| PEDOT:PSS | GFP | GFP | 2.3 x $10^{-14}$ M | y =1.065 + 0.078*x*, $R^2$=0.99 | 11 % |
| | | mCherry | -- | -- | 4 % |
| | | Lysozyme | -- | -- | 4 % |
| | SARS-CoV-1 | SARS-1 RBD | 1.5 x $10^{-15}$ M | y =0.616 +0.037*x*, $R^2$=0.89 | 9 % |
| | | SARS-2 RBD | 2.4 x $10^{-14}$ M | y =0.317 +0.015*x*, $R^2$=0.962 | 5 % |
| | | SARS-2 Spike | 2.8 x $10^{-16}$ M | y =1.406 +0.073*x*, $R^2$=0.995 | 14 % |
| | | GFP | -- | -- | 9 % |
| p(g0T2-g6T2) | GFP | GFP | 1.4 x $10^{-17}$ M | y =1.381 +0.076*x*, $R^2$=0.94 | 9 % |
| | | mCherry | -- | -- | 3 % |
| | | Lysozyme | -- | -- | 3 % |
| | SARS-CoV-1 | SARS-1 RBD | 1.6 x $10^{-17}$ M | y =1.372 +0.077*x*, $R^2$=0.88 | 9 % |
| | | SARS-2 RBD | 4.8 x $10^{-14}$ M | y =1.477 +0.105*x*, $R^2$=0.93 | 12 % |
| | | SARS-2 Spike | 1.8 x $10^{-20}$ M | y =1.678 +0.081*x*, $R^2$=0.98 | 7 % |
| | | GFP | -- | -- | 9 % |
| | | | | **Sensors in Saliva** | |
| PEDOT:PSS | MERS-CoV | MERS-CoV Spike | 6.1 x $10^{-19}$ M | y =0.666 +0.034*x*, $R^2$=0.959 | 5 % |
| p(g0T2-g6T2) | SARS-CoV-1 | SARS-1 RBD | -- | -- | -- |
| | | SARS-2 RBD | 2.3 x $10^{-14}$ M | y =1.28 +0.066*x*, $R^2$=0.91 | 11 % |
| | | SARS-2 Spike | 1.2 x $10^{-21}$ M | y =1.409 +0.065*x*, $R^2$=0.98 | 10 % |
| | | GFP | -- | -- | 2 % |
| | MERS-CoV | MERS-CoV Spike | 5.7 x $10^{-19}$ M | y =1.914 +0.102*x*, $R^2$=0.993 | 6 % |

### Table S2. Protein constructs

| Construct Name | Description | Molecular weight (kDa) | Isoelectric Point | E(280 nm) $M^{-1}\,cm^{-1}$ | E(485 nm) $M^{-1}\,cm^{-1}$ | E(587 nm) $M^{-1}\,cm^{-1}$ |
|---|---|---|---|---|---|---|
| Peptide | MCA-SpyTag | 1.76 | 8.24 | 1,490 | - | - |
| GFP Nanobody | GFPnanobody-8aa-SpyCatcher-His10 | 28.39 | 5.81 | 38,390 | - | - |
| SARS-CoV nanobody (VHH72) | SARS 1 / 2 nanobody(VHH72)-SpyCatcher-3C-His8 | 29.05 | 5.2 | 46,870 | - | - |
| MERS-CoV nanobody (VHH04) | MERS nanobody(VHH04)-SpyCatcher-3C-His10 | 28.90 | 5.77 | 41,370 | - | - |
| Target (msfGFP) | snoopTag-msfGFP-3C-TwinStrep | 33.59 | 6.06 | 29,910 | 83,300 | |
| Negative control (mCherry) | FRB-24-mCherry-Wp3-TwinStrep | 42.91 | 6.1 | 74,830 | | 60,000 |
| SARS-CoV-1 RBD | (Sino Biological 40150-V08B2)<br><br>Spike Receptor Binding Domain residues R306 - F527 His tagged | 26.51 | 8.51 | 40,340 | | |
| SARS-CoV-2 RBD | (Sino Biological 40592-V08H)<br><br>Spike Receptor Binding Domain residues R319 - F541 His tagged | 26.5 | 8.9 | 33,350 | | |
| SARS-CoV-2 S1 | (Sino Biological 40591-V08B1)<br><br>Spike Protein S1 subunit residues 1 - 685(R) His tagged | 76.45 | 8.28 | 90,650 | | |
| MERS-CoV S1 | (Sino Biological 40069-V08H)<br><br>Spike Protein S1 subunit residues 1 - 725 His tagged | 79.9 | 6.19 | 100,510 | | |

### Table S3. Protein sequences

| Construct Name | Description | Sequence |
|---|---|---|
| Peptide | MCA-SpyTag | SGSGAHIVMVDAYKPTK |
| GFP nanobody | GFPnanobody-8aa-SpyCatcher-3C-His10 | MASQVQLVESGGALVQPGGSLRLSCAASGFPVNRYSMRWYRQAPGKER<br>EWVAGMSSAGDRSSYEDSVKGRFTISRDDARNTVYLQMNSLKPEDTAV<br>YYCNVNVGFEYWGQGTQVTVSSKGSGSGSGSVDTLSGLSSEQGQSGDM<br>TIEEDSATHIKFSKRDEDGKELAGATMELRDSSGKTISTWISDGQVKDFY<br>LYPGKYTFVETAAPDGYEVATAITFTVNEQGQVTVNGKATKGDAHISGL<br>EVLFQGPTGHHHHHHHHHH |
| SARS-CoV nanobody<br><br>(VHH72) | SARS 1 / 2 anti-RBD nanobody(VHH72)-SpyCatcher-3C-His8 | MTGQVQLQESGGGLVQAGGSLRLSCAASGRTFSEYAMGWFRQAPGKER<br>EFVATISWSGGSTYYTDSVKGRFTISRDNAKNTVYLQMNSLKPDDTAVY<br>YCAAAGLGTVVSEWDYDYDYWGQGTQVTVSSGSGSGSGSGSVDTLSGL<br>SSEQGQSGDMTIEEDSATHIKFSKRDEDGKELAGATMELRDSSGKTISTW<br>ISDGQVKDFYLYPGKYTFVETAAPDGYEVATAITFTVNEQGQVTVNGKA<br>TKGDAHISGLEVLFQGPTGHHHHHHHH |
| MERS-CoV nanobody<br><br>(VHH04) | MERS anti-RBD nanobody(VHH04)-SpyCatcher-3C-His10 | MTGEVQLQESGGGSVQAGGSLRLSCEASGTISSMYCMGWFRQAPGKER<br>EGVALFNRSTGVEYYRASVKGRFTISHDNAKNTVYLQMNSLKLEDTAV<br>YYCAAGPTCGGWYPGLYNYWGQGTQVTVSSGSGSGSGSVDTLSGLSSE<br>QGQSGDMTIEEDSATHIKFSKRDEDGKELAGATMELRDSSGKTISTWISD<br>GQVKDFYLYPGKYTFVETAAPDGYEVATAITFTVNEQGQVTVNGKATK<br>GDAHISGLEVLFQGPTGHHHHHHHHHH |
| Target (msfGFP) | snoopTag-msfGFP-3C-TwinStrep | MASKLGDIEFIKVNKGSGSGSGSVSKGEELFTGVVPILVELDGDVNGHKF<br>SVRGEGEGDATNGKLTLKFICTTGKLPVPWPTLVTTLTYGVQCFSRYPDH<br>MKQHDFFKSAMPEGYVQERTISFKDDGTYKTRAEVKFEGDTLVNRIELK<br>GIDFKEDGNILGHKLEYNFNSHNVYITADKQKNGIKANFKIRHNVEDGSV<br>QLADHYQQNTPIGDGPVLLPDNHYLSTQSKLSKDPNEKRDHMVLLEFVT<br>AAGITHGMDELYKTGSTGLEVLFQGPTGSAWSHPQFEKGTGSGTGSGTG<br>SWSHPQFEK |
| Negative control (mCherry) | FRB-24-mCherry-Wp3-TwinStrep | MTGILWHEMWHEGLEEASRLYFGERNVKGMFEVLEPLHAMMERGPQT<br>LKETSFNQAYGRDLMEAQEWCRKYMKSGNVKDLLQAWDLYYHVFRRI<br>SKTGGSGSGSGSGSGSGSGSGSGSTGVSKGEEDNSAIIKEFMRFKVHMEG<br>SVNGHEFEIEGEGEGRPYEGTQTAKLKVTKGGPLPFAWDILSPQFMYGSK<br>AYVKHPADIPDYLKLSFPEGFKWERVMNFEDGGVVTVTQDSSLQDGEFI<br>YKVKLRGTNFPSDGPVMQKKTMGWEASSERMYPEDGALKGEIKQRLKL<br>KDGGHYDAEVKTTYKAKKPVQLPGAYNVNIKLDITSHNEDYTIVEQYER<br>AEGRHSTGGSGPLPPYTSAWSHPQFEKGGGSGGGSGGGSWSHPQFEK |
